# Integrating etiological insights with machine learning for precision diagnosis of obstructive jaundice: Findings from a high-volume center

**DOI:** 10.1101/2024.07.15.24310411

**Authors:** Ningyuan Wen, Yaoqun Wang, Xianze Xiong, Jianrong Xu, Shaofeng Wang, Yuan Tian, Di Zeng, Xingyu Pu, Geng Liu, Bei Li, Jiong Lu, Nansheng Cheng

## Abstract

**Introduction:** Large-scale cohort studies exploring the etiology of obstructive jaundice (OJ) are scarce, with current serum-based diagnostic markers offering suboptimal performance. This study leverages the largest retrospective cohort of OJ patients to date to investigate its disease spectrum and to develop a novel diagnostic system.

**Methods:** This study involves two retrospective observational cohorts. The biliary surgery cohort (BS cohort, n=349) served for initial data exploration and external validation of ML models. The large general cohort (LG cohort, n=5726) enabled an in-depth analysis of etiologies and the determination of relevant diagnostic indicators, in addition to supporting ML model development. Interpretable ML techniques were employed to derive insights from the models.

**Results:** The LG cohort highlighted a diverse disease spectrum of OJ, including cholangiocarcinoma (10.39% distal, 10.01% perihilar, 5.59% intrahepatic), pancreatic adenocarcinoma (19.11%), and common bile duct stones (18.27%) as leading causes. Traditional serum markers such as CA 19-9 and CEA lacked standalone diagnostic accuracy. Two ML-based models (collectively termed the MOLT model) were developed: a classifier to differentiate benign from malignant causes (AUROC=0.862) and a multi-class model to further stratify malignant and benign diseases (ACC=0.777). Interpretable ML tools provided clarity on critical features, offering actionable insights and enhancing transparency in the decision-making process.

**Discussion:** This study elucidates the etiological spectrum of OJ, meanwhile providing a practical and interpretable ML-based diagnostic tool. By leveraging large-scale clinical data, our model provides a rapid and reliable primary assessment for patients with OJ, enabling clinicians to identify potential etiologies and guide further diagnostic workup.

**Highlights:** *What is known:* Currently, there is a deficit in large-scale cohort studies as well as practical diagnostic models for identifying the etiology of obstructive jaundice (OJ).

*What is new here:* Our study emerged as the largest cohort study regarding OJ to date, delineating the spectrum of diseases associated with this condition. Interpretable ML models based on common clinical laboratory tests were developed, collectively termed the MOLT model, which not only distinguishes between benign and malignant obstructions, but also further differentiates between calculous benign lesions, non-calculous benign lesions, metastatic malignancies, pancreato-biliary malignancies and other types of malignancies. These findings can support the identification of the underlying etiology of OJ in primary clinical settings, helping clinicians make well-informed decisions.

**Registration Number:** This is a retrospective cohort study, preregistered in Open Science Framework (registration DOI: https://doi.org/10.17605/OSF.IO/DC4B8).

## Introduction

Obstructive jaundice (OJ) is a common problem associated with various hepato-pancreato-biliary (HPB) diseases.^1^ Benign causes of OJ encompass a wide range of diseases, such as common bile duct stones (CBDS), benign biliary strictures, Mirizzi syndrome, etc. Malignant causes typically involve tumors originating from the pancreas, bile ducts, or ampulla of Vater, including pancreatic adenocarcinoma, cholangiocarcinoma, ampullary carcinoma, and more.^2–4^ Additionally, intrahepatic mass involving the hepatic hilus and metastatic tumors from other sites may also lead to this condition.^5^ ^6^ These diverse etiologies require careful evaluation and management to determine the appropriate course of treatment. Over the past few decades, significant advancements have been made in the diagnostic approaches for patients presenting with OJ. Serum-based diagnostics, including liver function tests (LFTs) and tumor markers, such as carbohydrate antigen 19-9 (CA 19-9) and carcinoembryonic antigen (CEA), have greatly facilitated the initial assessment of OJ. Advancements in imaging technology have further enhanced diagnostic accuracy of OJ-associated HPB diseases.^7–9^ Despite these advancements, challenges persist in these diagnostic approaches. Serum-based markers may lack specificity and sensitivity, limiting their utility as standalone diagnostic tools.^10^ Meanwhile, imaging modalities may encounter limitations in differentiating benign from malignant etiologies or accurately characterizing the extent of disease involvement, especially when the lesions are small.^11^ Moreover, access to advanced imaging techniques may be limited under certain healthcare settings, hampering timely diagnosis and management. Core needle biopsy (CNB), while useful for tissue sampling, may be limited by potential sampling error, complications, or difficulty in accessing anatomically challenging lesions^12^ ^13^. Recently, some liquid biopsy techniques have shown promising potential in detecting biomarkers associated with OJ, but these methods remain largely experimental and have yet to transition fully into clinical practice^14–16^. Another major limitation is the lack of large-scale cohort studies, which hampers efforts to fully understand the disease spectrum underlying this condition.

Hence, harnessing the power of state-of-the-art machine learning (ML) methods, renowned for their ability to extract intricate patterns from complex datasets, we aimed to develop a robust diagnostic tool for patients presenting with OJ.^17^ ^18^ This tool, enabling the integration of diverse clinical laboratory tests to enhance accuracy and efficiency of diagnosis, can be easily implemented due to its straightforward and user-friendly nature. Our objective was for it to not only differentiate between benign and malignant OJ, but also to classify more specific etiologies based on this foundation. Its efficacy was validated in real-world settings using an external surgical cohort, with interpretable ML techniques employed to offer transparency and insights into the decision-making process.

## Methods

### Study Setting and Population

This study was conducted at West China Hospital of Sichuan University, a tertiary referral center and one of the largest medical institutions in Southwest China, serving a diverse population that includes both urban and rural patients. As a high-volume center, it receives referrals from across the region, providing a broad and representative sample of patients with obstructive jaundice (OJ).

The study protocol was approved by the Ethics Committee for Biomedical Research at West China Hospital of Sichuan University and was preregistered in the Open Science Framework (registration DOI: https://doi.org/10.17605/OSF.IO/DC4B8). The study workflow is visualized in the graphical abstract.

### Study Design and Cohorts

This study involved two retrospective observational cohorts from a single center:

1. The Biliary Surgery Cohort (BS Cohort): This cohort served as the dataset for initial data exploration and external validation of machine learning (ML) models. It included patients presenting with OJ who were admitted to the Department of Biliary Surgery between February 2022 and September 2023. After screening with predefined criteria (Supplementary Figure 1), 349 patients were included.
2. The Large General Cohort (LG Cohort): This cohort was utilized for comprehensive data analysis and ML model construction. Data were reviewed from all hospitalized patients presenting with OJ in the general hospital between January 2008 and January 2022. After screening according to predefined inclusion/exclusion criteria (Figure 1), 5,726 patients were included.

**Figure 1.**
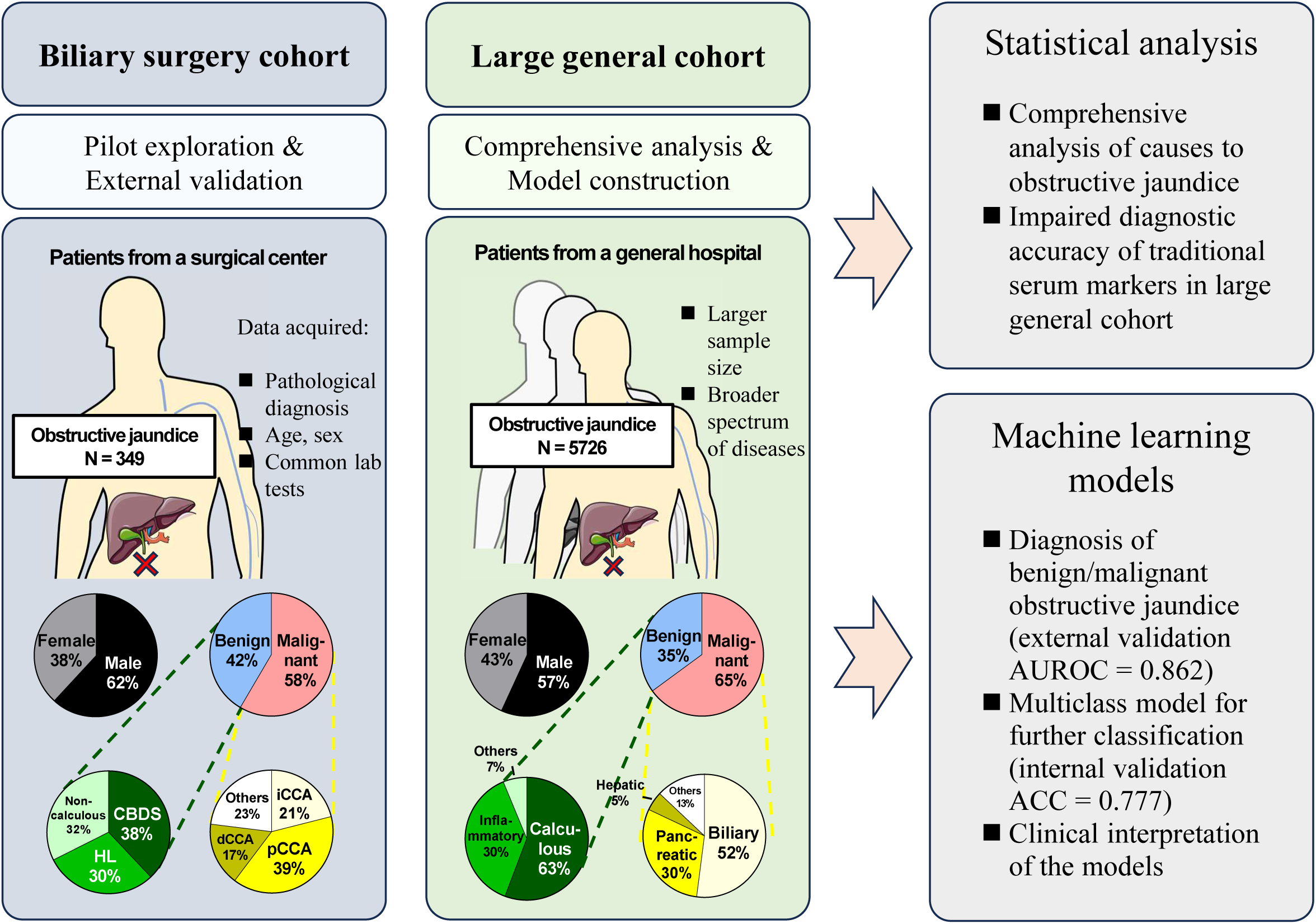
Graphical abstract of this study.

### Inclusion and Exclusion Criteria

The inclusion and exclusion criteria were designed to ensure a clear and consistent patient population for analysis. Inclusion criteria were as follows:

1. Documented clinical manifestation of OJ (regardless of whether it was listed as a primary or secondary diagnosis);
2. Reconfirmation of the diagnosis based on elevated cholestatic parameters (bilirubin, alkaline phosphatase, and γ-glutamyltransferase);
3. Etiology pathologically confirmed via ERCP, PTCD, CNB, or surgical intervention;
4. Age over 18 years.

Patients with OJ secondary to hepatopancreatobiliary (HPB) surgery were excluded to rule out iatrogenic factors. These criteria delineated the spectrum of diseases covered by this study, as depicted in Figure 2.

**Figure 2.**
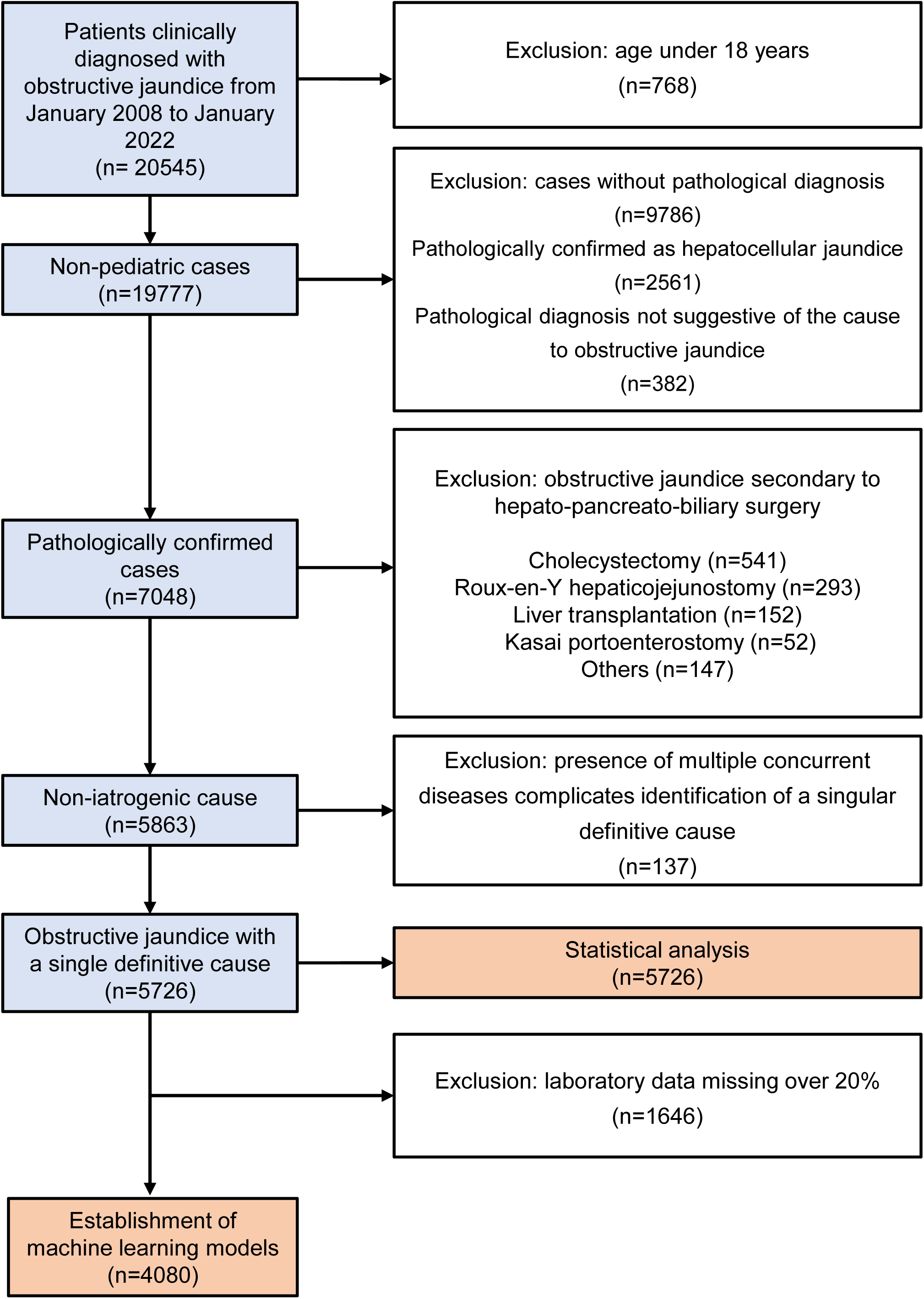
Flowchart illustrating the patient selection process from an initial pool of 20,545 patients to establish the LG cohort.

**Figure 3.**
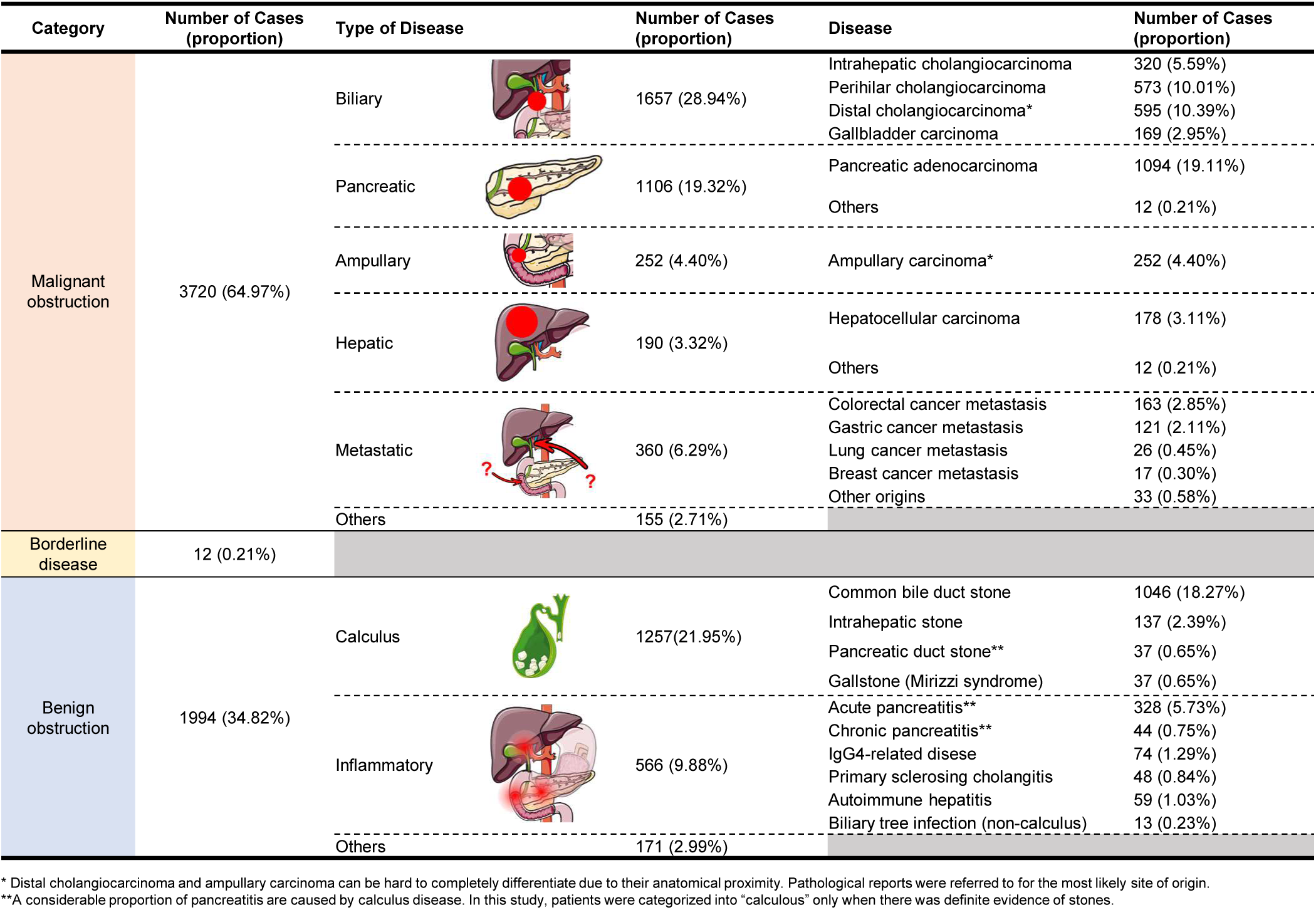
Summary chart depicting the disease spectrum of OJ observed in the LG cohort, highlighting the prevalence of various benign and malignant etiologies.

### Rationale for Single-Center Design

While the single-center design may limit generalizability, West China Hospital’s status as a high-volume referral center ensures a large and diverse patient population, including both primary and tertiary care cases. This setting provides a robust foundation for exploring the etiological spectrum of OJ and developing diagnostic models. However, we acknowledge that geographic and healthcare system differences may influence the applicability of our findings, and we have addressed this limitation in the Discussion section.

### Collection of clinical data

The clinical data of each patient was retrieved from the medical record archive of our institution and underwent de-identification. The following information underwent further investigation: age, sex, clinical diagnosis, pathological report and clinical laboratory test results. Given that the study was conducted at a single center in Southwest China, the majority of patients were of Han Chinese ethnicity, reflecting the regional population demographics. More detailed information on data processing is available in the Supplementary Materials.

### Development and validation of ML models

Based on these clinical data, we explored a diverse array of prediction models in two different tasks. The binary classification task focused on distinguishing between benign and malignant diseases, and the multi-class classification task intended to further categorize diseases into five detailed categories. Various ML algorithms were utilized to achieve best predictive performance, while internal validation and/or external validation were conducted for model evaluation. These models were meticulously interpreted. More detailed information on ML technique is available in the Supplementary Materials.

### Statistical analysis

Statistical analyses were conducted using R software. Shapiro-Wilk test and QQ plot were employed to assess the normality of data distribution. For comparisons between groups, independent samples t-tests and Mann-Whitney U tests were performed for continuous variables with and without normal distribution, respectively, while chi-square tests were conducted for categorical variables. The DeLong test was employed for the assessment of model performance. A significance level of P < 0.05 was considered statistically significant.

## Results

### Patient profiles and disease spectrums of the study cohorts

The biliary surgery cohort (BS cohort) included 349 consecutive surgical inpatients with OJ (Fig. S1). Demographically, there were 216 (62%) male and 133 (38%) female, with 60% of them aged over 60. In terms of the disease spectrum, there were 204 cases (58.5%) with malignant OJ and 145 cases (41.5%) with benign etiologies (Fig. S2A). As the BS cohort exclusively included patients from a biliary surgery center, these patients were predominantly associated with biliary malignancies, accounting for 49.0% of all types of diseases and 86.8% of all malignancies (Fig. S2C). Calculous diseases, including CBDS and hepatolithiasis (HL), accounted for 67.6% of all benign causes.

Undoubtedly, these preliminary results may not accurately reflect the true distribution of diseases in the general population, given that patients were selectively admitted to the surgical ward. Therefore, A large general cohort (LG cohort) comprising 5726 patients from a comprehensive medical center over a span of 14 years was analyzed (Fig. 1). Similarities were observed between the LG and BS cohorts in terms of sex, age, and the relative proportion of benign and malignant diseases, underscoring the representativeness of our previous observations (Fig. S2B). Still, statistical analysis revealed significant differences in the disease spectrum as well as other baseline characteristics between the two cohorts (Supplementary Table 1). Based on a larger sample size, the LG cohort was able to unveil some previously undisclosed insights into OJ. To sum up, biliary malignancies (1657 cases, 28.94%), pancreatic malignancies (1106 cases, 19.32%), ampullary malignancies (252 cases, 4.40%), hepatic malignancies (190 cases, 3.32%), metastatic cancers (360 cases, 6.29%) and other rare malignancies (155 cases, 2.71%) built up the malignant side of the disease spectrum; while calculous diseases (1257 cases, 21.95%), inflammatory diseases (566 case, 9.88%) and other benign causes (171 cases, 2.99%) composed the benign counterpart (Fig. 2). The top five leading causes of OJ were revealed as pancreatic adenocarcinoma (1094 cases, 19.11%), CBDS (1046 cases, 18.27%), distal cholangiocarcinoma (dCCA) (595 cases, 10.39%), perihilar cholangiocarcinoma (pCCA) (573 cases, 10.01%) and acute pancreatitis (non-calculous) (328 cases, 5.73%).

### Requirement for machine learning-based diagnostics in OJ

Our next objective was to ascertain whether a more effective diagnostic approach is warranted in distinguishing benign and malignant obstructions. A comparative analysis of the baseline characteristics between benign and malignant groups was conducted in the BS cohort and revealed a significant difference in multiple clinical indicators (Supplementary Table 2 and Supplementary Figure 3A). However, the diagnostic performance of single laboratory markers to distinguish benign from malignant conditions was suboptimal. The top five diagnostic markers were CA 19-9 (AUROC=0.768), DBIL (AUROC=0.736), TBIL (AUROC=0.730), CEA (AUROC=0.697), and DBIL/TBIL ratio (AUROC=0.696) (Supplementary Figure 3B).

The LG cohort was analyzed to validate these results based on a larger sample size. Comparative analysis unveiled statistical variances in the baseline characteristics between benign and malignant groups (Table 1).

**Table 1.** Comparative analysis of baseline characteristics between benign and malignant groups in the LG cohort.

| Variables | LG cohort (borderline disease excluded, n=5714) |  | pVal |
| --- | --- | --- | --- |
|  | Benign obstruction (n=1994) | Malignant obstruction (n=3720) |  |
| **Sex** |  |  | 0.36 |
| Male | 1151 (57.7%) | 2099 (56.4%) |  |
| Female | 843 (42.3%) | 1621 (43.6%) |  |
| **Age (years)** | 53.9 (16.1) | 59.9 (11.7) | <0.01 |
| **CA 19-9 (U/ml)** | 31.43 [13.63, 84.61] | 132.64 [34.76, 306.48] | <0.01 |
| **CEA (ng/ml)** | 2.12 [1.35, 3.35] | 3.41 [2.07, 6.39] | <0.01 |
| **CA 125 (U/ml)** | 19.33 [11.61, 41.29] | 24.35 [14.15, 51.65] | <0.01 |
| **AFP (ng/ml)** | 2.64 [1.90, 3.75] | 2.92 [2.14, 4.23] | <0.01 |
| **TBIL (μmol/L)** | 53.0 [42.4, 81.7] | 54.8 [44.3, 119.4] | <0.01 |
| **DBIL (μmol/L)** | 21.3 [13.3, 51.1] | 24.7 [16.5, 88.1] | <0.01 |
| **IBIL (μmol/L)** | 9.2 [6.6, 13.2] | 8.0 [5.6, 11.9] | <0.01 |
| **ALT (IU/L)** | 42 [24, 73] | 39 [26, 63] | 0.20 |
| **AST (IU/L)** | 41 [27, 75] | 33 [24, 57] | 0.02 |
| **ALP (IU/L)** | 139 [87, 226] | 131 [82, 229] | 0.42 |
| **γ-GT (IU/L)** | 119 [37, 288] | 81 [27, 195] | <0.01 |
| **ALB (g/L)** | 38.5 (5.3) | 37.5 (5.8) | <0.01 |
| **Glo (g/L)** | 28.5 (6.0) | 27.2 (5.2) | <0.01 |
| **A/G** | 1.37 [1.17, 1.56] | 1.41 [1.23, 1.61] | 0.07 |
| **GLU (mmol/L)** | 5.74 [5.17, 6.68] | 5.83 [5.23, 6.86] | 0.36 |
| **UREA (mmol/L)** | 4.8 [3.9, 5.8] | 4.7 [3.9, 5.6] | 0.59 |
| **CREA (μmol/L)** | 55.48 [52.00, 63.98] | 53.27 [48.11, 61.00] | 0.04 |
| **CysC (mg/L)** | 0.95 [0.84, 1.08] | 0.94 [0.83, 1.06] | 0.24 |
| **UA (μmol/L)** | 228 (79) | 207 (89) | 0.03 |
| **TG (mmol/L)** | 1.33 [1.02, 1.72] | 1.37 [1.03, 1.80] | 0.38 |
| **CHOL (mmol/L)** | 4.20 [3.45, 5.01] | 4.26 [3.42, 5.30] | 0.25 |
| **HDL (mmol/L)** | 1.00 [0.61, 1.32] | 0.87 [0.41, 1.29] | <0.01 |
| LDL (mmol/L) | 2.16 (0.87) | 2.06 (0.87) | 0.06 |
| CK (IU/L) | 65 [44, 104] | 71 [47, 101] | 0.31 |
| LDH (IU/L) | 177 [155, 203] | 179 [157, 201] | 0.79 |
| HBDH (IU/L) | 137 [124, 159] | 138 [123, 156] | 0.78 |
| RBC (×10<sup>12</sup>/L) | 3.84 (0.79) | 3.58 (0.65) | <0.01 |
| HGB (g/L) | 115 (22) | 109 (19) | <0.01 |
| HCT (L/L) | 0.35 (0.07) | 0.33 (0.06) | <0.01 |
| MCV (fL) | 93.0 [89.5, 96.3] | 93.7 [90.2, 97.0] | <0.01 |
| MCHC (g/L) | 328 (11) | 327 (11) | 0.26 |
| MCH (pg) | 30.6 [29.3, 31.7] | 30.8 [29.6, 32.0] | <0.01 |
| RDW-CV (%) | 14.6 [13.5, 16.0] | 15.2 [14.2, 16.6] | <0.01 |
| RDW-SD (fL) | 49.1 [45.1, 53.5] | 51.6 [47.9, 55.9] | <0.01 |
| PLT (×10<sup>9</sup>/L) | 187 [138, 241] | 194 [148, 245] | <0.01 |
| MPV (fL) | 11.9 (1.2) | 11.8 (1.2) | 0.35 |
| PDW (%) | 15.3 (3.2) | 15.1 (3.1) | 0.08 |
| P-LCR% (%) | 39.9 (9.8) | 39.5 (9.9) | 0.27 |
| WBC (×10<sup>9</sup>/L) | 6.97 [5.54, 9.26] | 7.36 [5.74, 9.52] | <0.01 |
| NEUT% (%) | 72.0 [65.0, 78.9] | 73.6 [67.3, 79.3] | <0.01 |
| LYM% (%) | 17.3 [11.6, 23.9] | 15.0 [10.7, 20.9] | <0.01 |
| MONO% (%) | 7.0 [5.9, 8.3] | 7.2 [6.2, 8.5] | <0.01 |
| EO% (%) | 1.4 [0.8, 2.3] | 1.5 [0.9, 2.4] | 0.04 |
| BASO% (%) | 0.4 [0.3, 0.5] | 0.4 [0.3, 0.5] | 0.58 |
| NEUT# (×10<sup>9</sup>/L) | 4.81 [3.58, 6.95] | 5.27 [3.87, 7.44] | <0.01 |
| LYM# (×10<sup>9</sup>/L) | 1.17 [0.85, 1.53] | 1.09 [0.83, 1.41] | <0.01 |
| MONO# (×10<sup>9</sup>/L) | 0.48 [0.38, 0.62] | 0.52 [0.41, 0.66] | <0.01 |
| EO# (×10<sup>9</sup>/L) | 0.09 [0.05, 0.16] | 0.10 [0.06, 0.16] | <0.01 |
| BASO# (×10<sup>9</sup>/L) | 0.03 [0.02, 0.04] | 0.03 [0.02, 0.04] | 0.26 |
| PT (s) | 11.7 [10.9, 12.8] | 11.6 [11.0, 12.6] | 0.38 |
| INR | 1.04 [0.97, 1.14] | 1.03 [0.97, 1.12] | 0.06 |
| **APTT (s)** | 28.1 [26.1, 30.8] | 28.0 [26.1, 30.8] | 0.85 |
| **Fbg (g/L)** | 3.45 (1.24) | 3.66 (1.08) | <0.01 |
| **TT (s)** | 18.4 [17.3, 19.5] | 18.2 [17.1, 19.4] | 0.03 |
| **CRP (mg/L)** | 14.8 [4.7, 42.4] | 19.8 [6.8, 72.5] | <0.01 |

Continuous variables with normal distribution are presented as mean value (SD) while others are presented as median (IQR). Categorical variables are presented as frequency (percentage).

A greater number of laboratory indicators were identified to show statistically significant distinctions in the LG cohort as opposed to the BS cohort. In contrast, the diagnostic efficacy of tumor markers in the LG cohort was generally less satisfactory. (Figure 4A). Among the top five indicators, only CA 19-9 had an AUROC exceeding 0.7 (AUROC=0.712), followed by CEA (AUROC=0.685), Age (AUROC=0.617), RDW-SD (AUROC=0.616) and DBIL (AUROC=0.613) (Figure 4B). The DeLong’s test indicated a significant decrease in diagnostic efficacy of indicators including CA 19-9 and DBIL in the LG cohort (Figure 4C). These results suggested that in a larger, more comprehensive patient cohort with a more complex diagnostic environment, individual serum markers are not robust diagnostic tools for patients presenting with OJ. Hence, there is a necessity to build a more efficient diagnostic tool.

**Figure 4.**
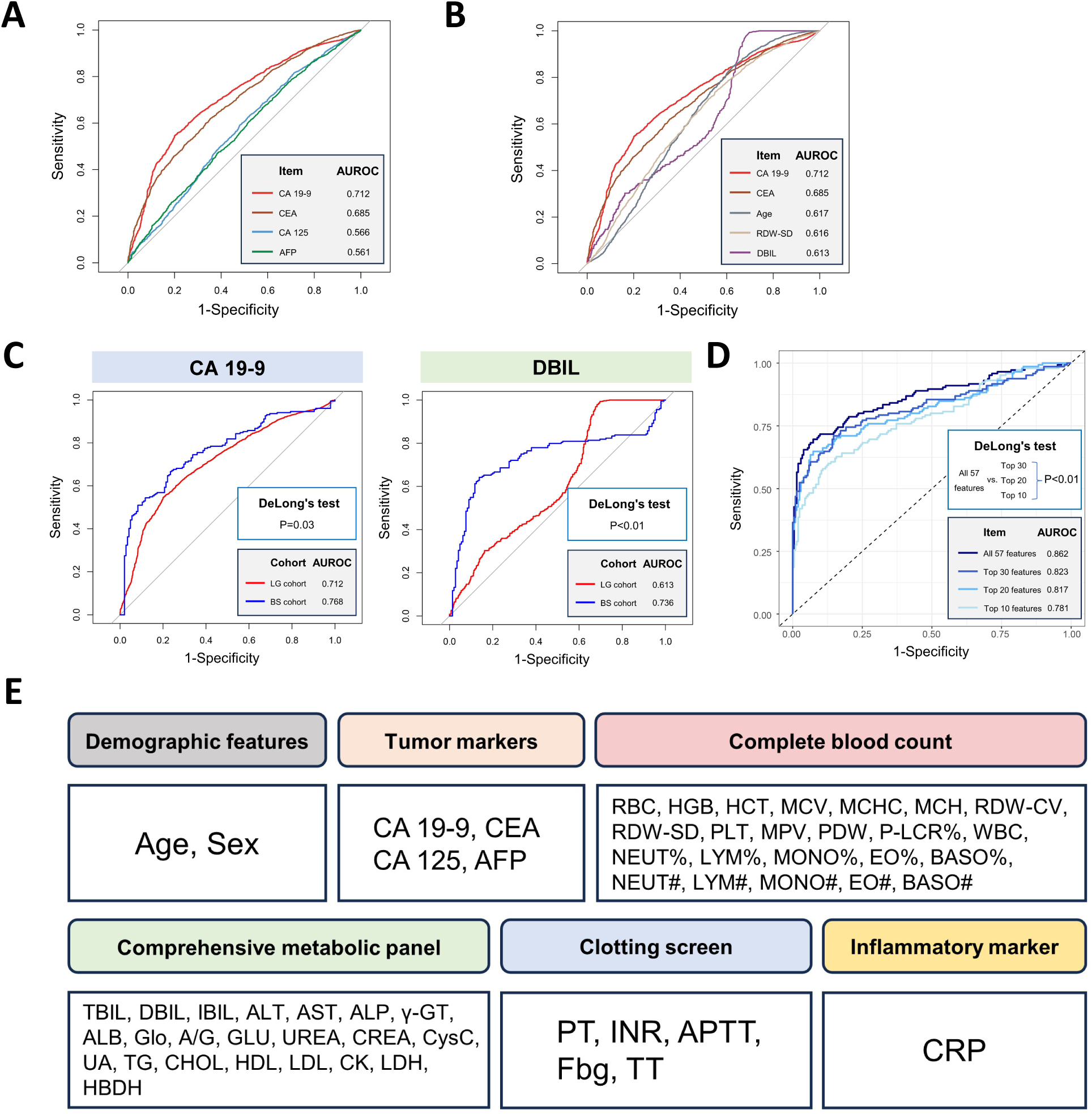
The necessity and feasibility of constructing a ML model for the clinical diagnosis of OJ. Traditional serum makers, including **(A)** tumor makers and **(B)** top 5 biomarkers ranked by AUROC, were found to exhibit suboptimal diagnostic efficacy in the LG cohort. **(C)** In the LG cohort, the diagnostic scenario becomes more intricate, causing markers like CA 19-9 and DBIL to exhibit diminished efficacy compared to the BS cohort, necessitating the development of combined ML models. **(D)** The number of features included in the ML model significantly influenced its diagnostic efficacy, as ML model with 57 features outperformed the others (DeLong’s test p < 0.01). **(E)** The included features encompass a wide spectrum of clinical characteristics including demographic features, tumor markers, complete blood count, comprehensive metabolic panel, clotting screen and inflammatory markers.

We first attempted a traditional linear regression model to integrate the diagnostic power of multiple test indicators. The diagnostic model construction process, employing stepwise logistic regression, incorporated a selection procedure of 57 variables (Supplementary Figure 4A). The diagnostic efficacy of this model was found to be moderate, achieving AUROC values of 0.784 and 0.791 in the internal and external validation sets, respectively. Notably, when compared to the subsequently established ML model, it presented significantly lower AUROC values as well as inadequate sensitivity and specificity (Supplementary Figure 4B & 4C). Similarly, in subsequently established ML models, the number of features included in the model significantly influenced its diagnostic efficacy (Figure 4D). These results highlighted the inherent advantage of ML techniques in leveraging the diagnostic power of multiple parameters or features. Therefore, the subsequently established ML models included all 57 features delineating common dimensions of disease characteristics (Figure 4E).

### Establishment, validation and interpretation of binary MOLT model to distinguish benign and malignant obstructions

After confirming the feasibility of constructing a ML diagnostic model based on 57 common clinical features, we proceeded to optimize its diagnostic performance. A series of mainstream ML methods were employed to construct this model, with their performance compared to select the optimal one. DeLong’s test between ROC curves was conducted to finalize our choice (Supplementary Figure 5). The RF model ultimately stood out for its remarkable performance both in the internal and external validation sets (Figure 5A-C and Table 2).

**Figure 5.**
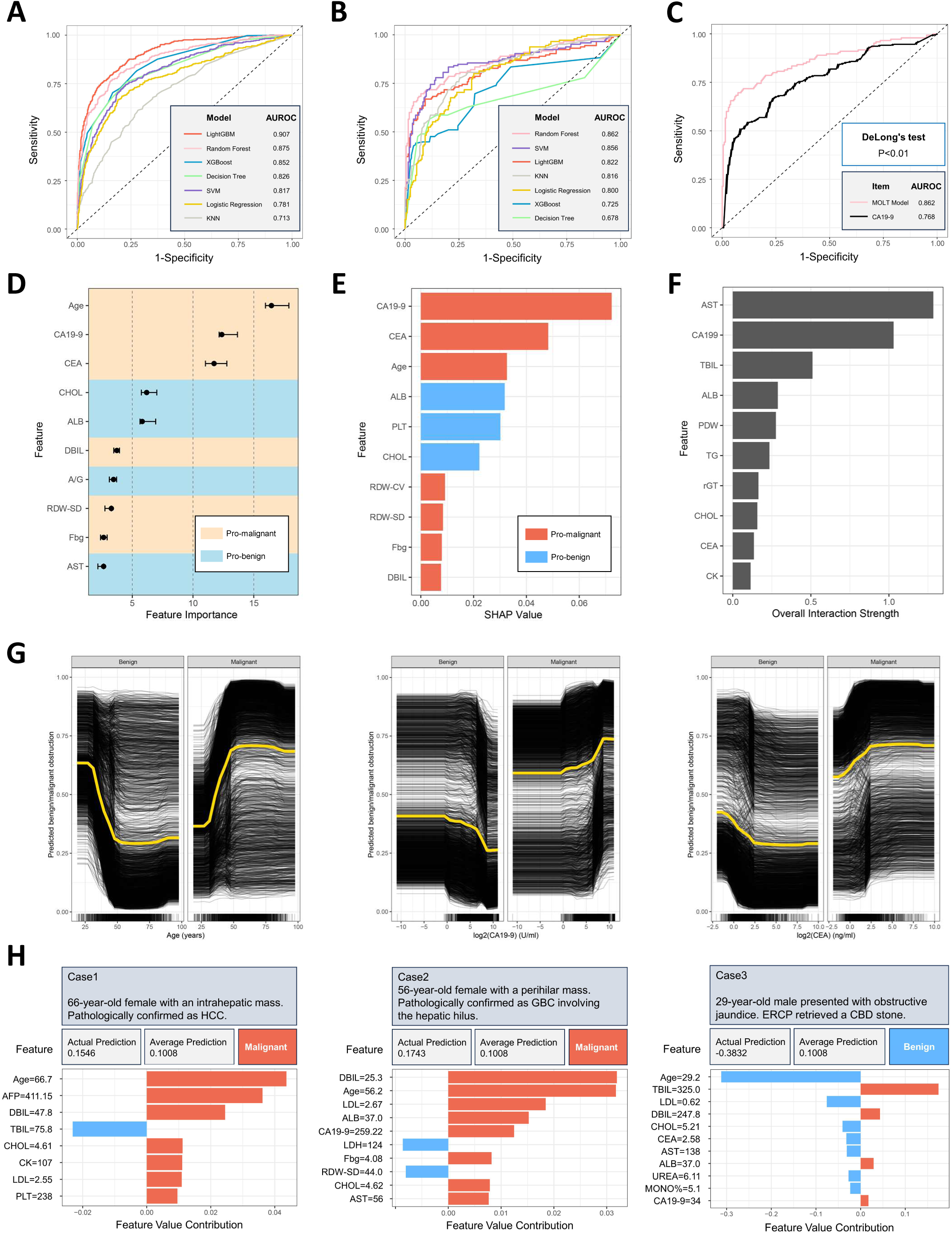
The establishment, validation and interpretation of binary MOLT model. **(A)** In the internal validation set, the lightGBM model showcased best performance measured by AUROC, while **(B)** the RF model showcased best performance in the external validation set. **(C)** The RF model was subsequently designated as the MOLT model, which exhibited better performance compared with traditional CA 19-9 in the external validation set. Interpretation of the MOLT model revealed **(D)** features with top-ranked feature importance score, **(E)** features with top-ranked SHAP values and **(F)** features with top-ranked interaction score. **(G)** PDP and ICE plots were created for the top three features (age, CA 19-9, and CEA) identified by SHAP values, elucidating their impact on the model’s predictions. **(H)** The SHAP-interpreted ML model clarified individualized decision-making processes, offering understanding into prediction rationale for each case.

**Table 2.** Comparison of performance among various ML methods employed for construction of the binary MOLT model.

| Cohort | Model | Index |  |  |  |  |  |
| --- | --- | --- | --- | --- | --- | --- | --- |
|  |  | AUROC (95%CI) | ACC | AUPR | F1 score | Sensitivity | Specificity |
| **LG cohort**<br>**(training)** | Random Forest | 1.000 (1.000-1.000) | 0.996 | 1.000 | 0.997 | 0.987 | 1.000 |
|  | LightGBM | 0.999 (0.999-1.000) | 0.983 | 0.999 | 0.988 | 0.956 | 0.996 |
|  | XGBoost | 0.891 (0.878-0.904) | 0.836 | 0.842 | 0.888 | 0.573 | 0.961 |
|  | Decision Tree | 0.836 (0.820-0.853) | 0.825 | 0.744 | 0.874 | 0.681 | 0.894 |
|  | SVM | 0.920 (0.907-0.932) | 0.872 | 0.892 | 0.911 | 0.680 | 0.963 |
|  | Logistic Regression | 0.811 (0.793-0.829) | 0.787 | 0.707 | 0.855 | 0.501 | 0.923 |
|  | KNN | 0.992 (0.990-0.995) | 0.907 | 0.982 | 0.935 | 0.731 | 0.990 |
| **LG cohort**<br>**(internal validation)** | Random Forest | 0.875 (0.855-0.896) | 0.824 | 0.816 | 0.878 | 0.566 | 0.955 |
|  | LightGBM | 0.907 (0.891-0.923) | 0.842 | 0.851 | 0.887 | 0.650 | 0.938 |
|  | XGBoost | 0.852 (0.832-0.873) | 0.798 | 0.774 | 0.862 | 0.504 | 0.947 |
|  | Decision Tree | 0.826 (0.803-0.849) | 0.791 | 0.748 | 0.848 | 0.613 | 0.881 |
|  | SVM | 0.817 (0.793-0.842) | 0.772 | 0.719 | 0.841 | 0.500 | 0.910 |
|  | Logistic Regression | 0.781 (0.754-0.809) | 0.757 | 0.672 | 0.835 | 0.431 | 0.923 |
|  | KNN | 0.713 (0.684-0.743) | 0.705 | 0.582 | 0.799 | 0.354 | 0.883 |
| **BS cohort**<br>**(external validation)** | Random Forest | 0.862 (0.819-0.904) | 0.828 | 0.865 | 0.864 | 0.676 | 0.936 |
|  | LightGBM | 0.822 (0.774-0.870) | 0.802 | 0.827 | 0.841 | 0.669 | 0.897 |
|  | XGBoost | 0.725 (0.667-0.782) | 0.699 | 0.709 | 0.768 | 0.483 | 0.853 |
|  | Decision Tree | 0.678 (0.615-0.741) | 0.731 | 0.689 | 0.783 | 0.586 | 0.833 |
|  | SVM | 0.856 (0.812-0.900) | 0.808 | 0.844 | 0.828 | 0.834 | 0.789 |
|  | Logistic Regression | 0.800 (0.754-0.846) | 0.722 | 0.734 | 0.771 | 0.614 | 0.799 |
|  | KNN | 0.816 (0.770-0.862) | 0.751 | 0.781 | 0.806 | 0.559 | 0.887 |

Cells labeled red/yellow represent the first/second-best performance in each cohort.

Subsequently, the RF model was selected and designated as the binary MOLT model, standing for **M**achine learning of **O**bstructive jaundice based on common **L**aboratory **T**ests.

To present the decision-making process of the binary MOLT model in a more transparent manner, we employed several methods for model interpretation. Feature importance scores highlighted the top-ranking features distinguishing benign from malignant etiologies. The top 10 features were identified as age, CA 19-9, CEA, CHOL, ALB, DBIL, A/G, RDW-SD, Fbg and AST (Figure 5D and Supplementary Figure 6A). While feature importance scores provide a global view of feature importance across the dataset, SHAP values offer a more nuanced understanding of how each feature influences individual predictions, taking interactions and dependencies between features into account ^19^ ^20^. Top 10 features ranked by SHAP value were CA 19-9, CEA, age, ALB, PLT, CHOL, RDW-CV, RDW-SD, Fbg and DBIL (Figure 5E and Supplementary Figure 6B). The overall interaction strength provided insights into the complexity of relationships between predictor variables in the binary MOLT model (Figure 5F). Partial Dependence Plots (PDP) and Individual Conditional Expectation (ICE) Plots were generated for the top three features identified by SHAP values (age, CA 19-9 and CEA), illustrating how individual feature values impact the model’s predictions (Figure 5G). With the SHAP-interpreted ML model, individualized decision-making processes were elucidated, allowing for a comprehensive understanding of how the model arrives at predictions for each specific case (Figure 5H). Decision boundaries pertaining to key features were also visualized (Supplementary Figure 7).

### Establishment, validation and interpretation of multi-class MOLT model for further classification

Building upon the initial model, we converted the binary classification target into a multi-classification target to construct a more complex model, namely the multi-class MOLT model, which differentiates between calculous benign lesions, non-calculous benign lesions, metastatic malignancies, pancreato-biliary malignancies and other types of malignancies (Figure 6A). Similar to the original binary MOLT model, a series of ML methods were employed, with the best-performing one selected to optimize model performance (Figure 6B). Notably, external validation was unable to be carried out for the multi-class MOLT model as there were no metastatic patient in the BS cohort. Outperforming the others, the XGBoost model showcased impressive diagnostic efficiency, boasting an ACC of 0.777 and an AUNU of 0.882, a notable achievement given the complexity of the task encompassing five classes. ML interpretability tools were also utilized to explain the multi-class MOLT model. Feature importance scores highlighted the top-ranking features contributing to this model, namely CA 19-9, age, CEA, ALB, CHOL, AFP, UA, A/G, RBC and CA125 (Figure 6C). SHAP values were utilized to assess how individual features influenced the model’s decisions within each disease category (Figure 6D). ALB, A/G, CA 19-9, CEA and AST were top 5 features contributing to the diagnosis of benign calculous disease; CA19-9, PDW, ALB, CEA and P-LCR% were top 5 features contributing to the diagnosis of benign non-calculous disease; CEA, age, TBIL, CA 19-9 and RBC were top 5 features contributing to the diagnosis of metastatic malignancies; ALB, CA 19-9, CEA, A/G and RBC were top 5 features contributing to the diagnosis of pancreato-biliary malignancies; while AFP, PLT, ALB, A/G and HDL were top 5 features contributing to the diagnosis of other malignancies.

**Figure 6.**
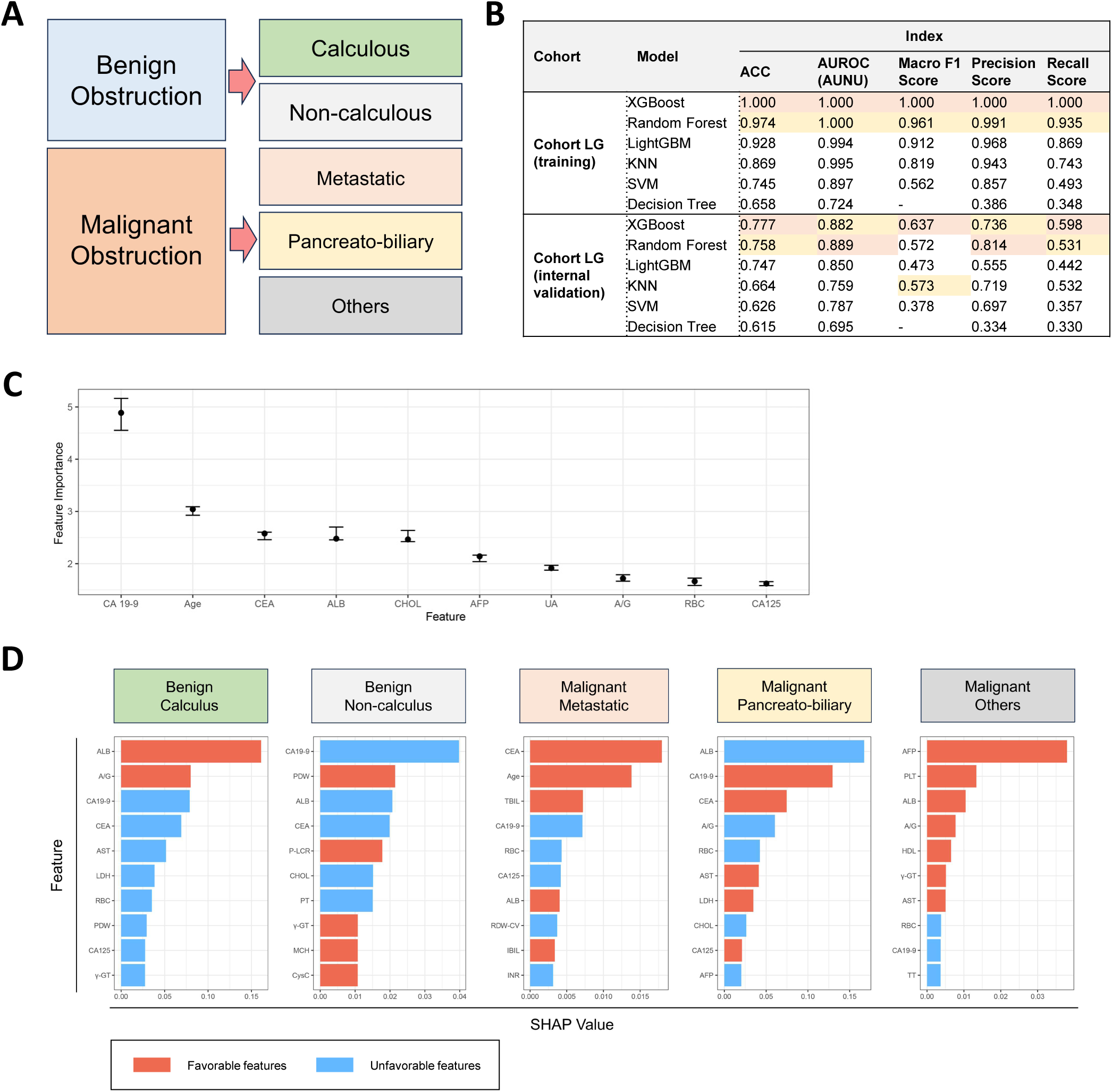
The establishment, validation and interpretation of multi-class MOLT model. **(A)** Extending the binary MOLT model, a five-class ML task was formulated. **(B)** The performance of diverse multi-class models was gauged using metrics like ACC, AUNU, macro F1 score, precision and recall scores. Similarly, the decision-making process of the multi-class MOLT model was elucidated with **(C)** feature importance score and **(D)** SHAP value.

## Discussion

Until now, large-scale cohorts regarding obstructive jaundice (OJ) remain scarce, which has led to a limited understanding of its disease spectrum and the efficacy of existing diagnostic approaches. Our study included the largest retrospective cohort of OJ to-date, providing valuable insights into its disease spectrum and lab test data through comprehensive analysis. Consequently, we developed machine learning (ML) models based on common clinical laboratory tests, which not only distinguishes between benign and malignant obstructions, but also further differentiates between calculous benign lesions, non-calculous benign lesions, metastatic malignancies, pancreato-biliary malignancies and other types of malignancies. To ensure transparency in the decision-making process, interpretable ML tools were utilized to decipher these models.

At first, it is important to address the extent to which this diagnostic model can aid clinical decision-making and ultimately improve patient outcomes. In our view, the MOLT model is not intended to replace imaging or interventional procedures, such as MRCP or ERCP, which remain essential for definitive diagnosis and treatment. Instead, the primary value of our model lies in its ability to provide a rapid, non-invasive, and interpretable “primary assessment” or “quick assessment” for patients with OJ. By leveraging commonly available laboratory indices, the MOLT model can help clinicians prioritize potential etiologies early in the diagnostic process. This can guide the selection of subsequent diagnostic tests (e.g., diverse imaging modalities) and streamline the diagnostic pathway, potentially reducing delays in diagnosis and treatment initiation. For instance, in resource-limited settings or for patients who cannot immediately undergo advanced imaging, our model could serve as a valuable triage tool. This approach aligns with strategies previously proposed for colorectal cancer diagnosis and other public health initiatives^21^ ^22^.

Before establishing the model, the disease spectrum of OJ needs to be carefully examined, as it directly impacts diagnosis, treatment, and healthcare resource allocation. Undoubtedly, the disease spectrum of OJ may vary across different countries and regions. However, the dearth of knowledge in this field has restricted our understanding in this field. Our study emerged as a significant contribution by offering a comprehensive analysis of over 5000 patients with OJ in a single Chinese center over a period of 14 years. Smaller retrospective cohort studies conducted across Europe, Australia, Central Asia and South Asia provided valuable insights into OJ.^23–29^ Notably, Garcea et al.’s retrospective analysis of over 1000 cases in the United Kingdom found similar disease patterns to ours, with CBD stones and pancreatic ductal adenocarcinoma as primary benign and malignant etiologies, respectively.^26^ Similarly, Björnsson et al.’s analysis of 241 patients in Sweden revealed a higher incidence of malignant obstruction (63.9%) compared to benign cases, with cholangiocarcinoma accounting for one-third of malignant obstructions, mirroring our findings.^23^ These findings suggest that the disease spectrum of OJ may be more consistent across different regions than previously believed. Moreover, our study supplemented these insights by revealing additional dimensions. Firstly, we observed that non-calculous benign etiologies might have been underreported, as most of the previous studies identified CBDS as the predominant benign cause. Our findings indicated that CBDS only accounted for approximately half of the benign cases, while around one-third were associated with diverse non-calculous factors. In addition, there may have been an underestimation of metastatic causes of OJ, considering the lower likelihood of obtaining a pathological diagnosis in these patients. Furthermore, intrahepatic lesions involving the hepatic hilus, mainly intrahepatic cholangiocarcinoma and hepatocellular carcinoma, constitute a significant proportion (approximately 10%) of all cases. Our study also yielded valuable insights in diagnostics. We not only presented a practical diagnostic tool for distinguishing between different causes to OJ, but also enhanced comprehension by providing transparency into the decision-making process. According to previous studies, calculous diseases in benign OJ can be accurately distinguished through various imaging modalities.^30–32^ However, distinguishing non-calculous benign diseases from malignant diseases can be considerably challenging ^33–36^. As our study revealed that over one-third of the benign cases were associated with non-calculous etiologies, there is a need to place greater emphasis on addressing this issue. Meanwhile, in the realm of malignancy, clinicians seek to stratify cancers according to their level of aggressiveness. To this end, we employed ML techniques to develop two models: one effectively distinguishes between benign and malignant causes, while the other offers nuanced insights by further classifying malignancies into three tiers and benign diseases into two. Subsequently, these models may facilitate the application of appropriate diagnostic and therapeutic interventions.

While we do not claim that our model can replace current diagnostic standards, we believe it complements existing methods by offering a cost-effective, accessible, and efficient preliminary assessment. This could be particularly beneficial in scenarios where rapid triage or resource optimization is critical. It is noteworthy that the concept of integrating diverse laboratory test results into a unified model did not arise arbitrarily. There has long been ample evidence pointing towards this direction. For instance, multiple studies have observed that patients with malignant obstruction tends to be older than the benign group.^23^ ^27^ Similarly, benign OJ is observed to be associated with lower bilirubin levels, as biliary obstructions caused by calculous disease tend to be intermittent.^1^ ^26^ Furthermore, there is documented evidence suggesting an association between OJ and renal injury, with the severity of renal dysfunction potentially reflecting the nature of the disease.^37–39^ In this study, by leveraging interpretable ML models, we gained further insight into how traditional and novel biomarkers contribute to the diagnostic model. While established markers such as CA 19-9, CEA, age, and bilirubin levels remained significant, factors like albumin levels, cholesterol levels, and red cell distribution width (RDW-SD) emerged as noteworthy contributors, offering new dimensions to the diagnostic evaluation of OJ.

CA 19-9, with a standalone AUROC of 0.712, demonstrated the highest predictive value among the top five indicators. This aligns with its well-established role as a tumor marker for pancreato-biliary malignancies, particularly pancreatic ductal adenocarcinoma and cholangiocarcinoma^40^ ^41^. However, its elevation in benign conditions, such as cholangitis or biliary obstruction, underscores the need for careful interpretation in clinical practice^10^ ^42^.

CEA, another well-established tumor marker, showed a standalone AUROC of 0.685. While its sensitivity and specificity for OJ are generally lower than those of CA 19-9, it remains a valuable adjunct in differentiating malignant from benign causes^43^ ^44^.

Age (standalone AUROC = 0.617) emerged as a significant predictor, consistent with the known association between advancing age and an increased risk of malignant biliary diseases. Older patients are more likely to present with malignancies such as pancreatic cancer or cholangiocarcinoma, whereas benign causes like choledocholithiasis are more prevalent in younger populations^45^.

Direct bilirubin (DBIL), with a standalone AUROC of 0.613, reflects the severity of biliary obstruction. Our findings suggest that DBIL levels are typically higher in malignant OJ, likely due to the more complete and persistent nature of obstruction in malignancies^46^. In contrast, benign OJ, often caused by conditions such as CBDS, may present with intermittent or partial obstruction, leading to relatively lower DBIL levels^47^.

Finally, RDW-SD (standalone AUROC = 0.616), though less conventional, represents an intriguing biomarker in the context of OJ. Traditionally associated with hematologic disorders, RDW-SD has been linked to systemic inflammation and liver dysfunction, particularly in decompensated liver cirrhosis^48^. Its inclusion in our model highlights the complex interplay between hepatic dysfunction and systemic inflammatory responses, warranting further investigation into its diagnostic and prognostic utility.

Several limitations of this study should be noted. Firstly, as this study exclusively enrolled patients with a confirmed pathological diagnosis, the findings regarding the proportions of specific types of diseases contributing to OJ may be subject to bias due to variations in the likelihood of different diseases to be biopsied. Secondly, this is a single-center study, primarily involving patients from China. As a result, the potential impact of geographic variations among patients and differences in detection methods across various clinical laboratories on the study outcomes was not addressed, even though we observed similarities in our findings with previous researches from other regions. Furthermore, our MOLT model exhibited exceptional specificity; however, its sensitivity fell short of expectations. This underscores the need for future studies to improve both the specificity and sensitivity of diagnostics. Despite these limitations, our study stands as the largest cohort study in the field of OJ to date, with robust diagnostic tools developed through the utilization of state-of-the-art techniques.

To conclude, our study developed the MOLT model for the diagnosis of patients presenting with OJ, which may facilitate personalized and user-friendly clinical decision-making of this condition. By providing a rapid, interpretable, and non-invasive primary assessment, our model complements existing diagnostic methods and has the potential to streamline clinical workflows, particularly in resource-limited settings or for patients requiring immediate triage.

## Data Availability

All data produced in the present study are available upon reasonable request to the authors.

https://doi.org/10.17605/OSF.IO/DC4B8

## Acknowledgements

We would like to express our gratitude to Professor Jingxin Zhang from Harbin University of Commerce for imparting ML methods based on the mlr3 framework in R. We would also like to express our appreciation to Smart Server Medical Art (https://smart.servier.com/) and Scidraw (https://scidraw.io/) for providing free medical illustrations.

## Specific author contributions

Study concept and design: NW, YW, BL, JL, GL and NC; Data acquisition: NW, YW, YT, BL, JL and NC; Data analysis and interpretation: NW, YW, GL, JX and DZ; Implementation of machine learning: NW, YW, XP and GL; Drafting of the manuscript: NW, SW, GL, BL, JL and CN; Funding: XX, BL, JL and CN. All authors have read and critically revised the manuscript and agreed to the published version.

## Data sharing

We have made the source code and datasets used in this study publicly available on GitHub. The project, titled “Machine Learning of Obstructive Jaundice based on Common Laboratory Tests (the MOLT model)”, can be accessed at https://github.com/re5yho/Machine-learning-of-Obstructive-jaundice-based-on-common-Laboratory-Tests-the-MOLT-model-.git. Researchers interested in replicating or extending our work are encouraged to explore the repository.

## Transparency statement

The primary author (NC, the manuscript’s guarantor) confirms that the manuscript provides a truthful, precise, and transparent portrayal of the study being presented. No crucial elements of the study have been excluded, and any deviations from the original study plan (and, if applicable, registration) have been elucidated.

## Patient and public involvement statement

Our statement of intent for patient and public involvement outlines our commitment to collaborating with individuals affected by obstructive jaundice to shape and guide our research projects aimed at advancing our understanding of this condition. We will also seek input from members of the public, especially in areas such as obstructive jaundice epidemiology, prevention, early detection and diagnosis.

## Abbreviations

ML: machine learning
ERCP: endoscopic retrograde cholangiopancreatography
PTCD: percutaneous transhepatic cholangiography drainage
iCCA: intrahepatic cholangiocarcinoma
pCCA: perihilar cholangiocarcinoma
dCCA: distal cholangiocarcinoma
AFP: α-fetoprotein
CEA: carcinoembryonic antigen
CA 125: cancer antigen 125
CA 19-9: cancer antigen 19-9
RBC: red blood cell count
HGB: hemoglobin
HCT: hematocrit
MCV: mean corpuscular volume
MCHC: mean corpuscular hemoglobin concentration
MCH: mean corpuscular hemoglobin
RDW-CV: red cell distribution width-coefficient of variation
RDW-SD: red cell distribution width-standard deviation
PLT: platelet count
MPV: mean platelet volume
PDW: platelet distribution width
P-LCR%: large platelet ratio
WBC: white blood cell count
NEUT%: neutrophil percentage
LYM%: lymphocyte percentage
MONO%: monocyte percentage
EO%: eosinophil percentage
BASO%: basophil percentage
NEUT#: neutrophil count
LYM#: lymphocyte count
MONO#: monocyte count
EO#: eosinophil count
BASO#: basophil count
TBIL: total bilirubin
DBIL: direct bilirubin
IBIL: indirect bilirubin
ALT: alanine aminotransferase
AST: aspartate aminotransferase
ALP: alkaline phosphatase
γ-GT: γ-glutamyl transferase
ALB: albumin
Glo: globulin
A/G: albumin/globulin ratio
GLU: glucose
UREA: urea
CREA: creatinine
CysC: cystatin C
UA: uric acid
TG: triglycerides
CHOL: cholesterol
HDL: high-density lipoprotein cholesterol
LDL: low-density lipoprotein cholesterol
CK: creatine kinase
LDH: lactate dehydrogenase
HBDH: hydroxybutyrate dehydrogenase
PT: prothrombin time
INR: international normalized ratio
APTT: activated partial thromboplastin time
Fbg: fibrinogen
TT: thrombin time
CRP: C-reactive protein.

## References

1. Jarnagin WR. Blumgart’s Surgery of the Liver, Biliary Tract and Pancreas, 2-Volume Set: Elsevier Health Sciences 2022.

2. Pereira SP, Goodchild G, Webster GJM. The endoscopist and malignant and non-malignant biliary obstruction. Biochim Biophys Acta Mol Basis Dis 2018;1864(4 Pt B):1478–83. doi: 10.1016/j.bbadis.2017.09.013 [published Online First: 2017/09/22]

3. Kapoor BS, Mauri G, Lorenz JM. Management of Biliary Strictures: State-of-the-Art Review. Radiology 2018;289(3):590–603. doi: 10.1148/radiol.2018172424 [published Online First: 2018/10/24]

4. Fry DE. Obstructive jaundice. Causes and surgical interventions. Postgrad Med 1988;84(5):217–22, 27, 30. doi: 10.1080/00325481.1988.11700446 [published Online First: 1988/10/01]

5. Okamoto T. Malignant biliary obstruction due to metastatic non-hepato-pancreato-biliary cancer. World J Gastroenterol 2022;28(10):985–1008. doi: 10.3748/wjg.v28.i10.985 [published Online First: 2022/04/19]

6. Lu J, Li B, Li FY, et al. Long-term outcome and prognostic factors of intrahepatic cholangiocarcinoma involving the hepatic hilus versus hilar cholangiocarcinoma after curative-intent resection: Should they be recognized as perihilar cholangiocarcinoma or differentiated? Eur J Surg Oncol 2019;45(11):2173–79. doi: 10.1016/j.ejso.2019.06.014 [published Online First: 2019/06/19]

7. She YM, Ge N. The value of endoscopic ultrasonography for differential diagnosis in obstructive jaundice of the distal common bile duct. Expert Rev Gastroenterol Hepatol 2022;16(7):653–64. doi: 10.1080/17474124.2022.2098111 [published Online First: 2022/07/07]

8. Heilmaier C, Lutz AM, Bolog N, et al. Focal liver lesions: detection and characterization at double-contrast liver MR Imaging with ferucarbotran and gadobutrol versus single-contrast liver MR imaging. Radiology 2009;253(3):724–33. doi: 10.1148/radiol.2533090161 [published Online First: 2009/10/01]

9. Lv WJ, Zhao XY, Hu DD, et al. Insight into Bile Duct Reaction to Obstruction from a Three-dimensional Perspective Using ex Vivo Phase-Contrast CT. Radiology 2021;299(3):597–610. doi: 10.1148/radiol.2021203967 [published Online First: 2021/04/21]

10. Marrelli D, Caruso S, Pedrazzani C, et al. CA19-9 serum levels in obstructive jaundice: clinical value in benign and malignant conditions. Am J Surg 2009;198(3):333–9. doi: 10.1016/j.amjsurg.2008.12.031 [published Online First: 2009/04/21]

11. Joo I, Lee JM, Yoon JH. Imaging Diagnosis of Intrahepatic and Perihilar Cholangiocarcinoma: Recent Advances and Challenges. Radiology 2018;288(1):7–13. doi: 10.1148/radiol.2018171187 [published Online First: 2018/06/06]

12. Eschrich J, Kobus Z, Geisel D, et al. The Diagnostic Approach towards Combined Hepatocellular-Cholangiocarcinoma-State of the Art and Future Perspectives. Cancers (Basel) 2023;15(1) doi: 10.3390/cancers15010301 [published Online First: 2023/01/09]

13. Jakob J, Salameh R, Wichmann D, et al. Needle tract seeding and abdominal recurrence following pre-treatment biopsy of gastrointestinal stromal tumors (GIST): results of a systematic review. BMC Surg 2022;22(1):202. doi: 10.1186/s12893-022-01648-2 [published Online First: 2022/05/22]

14. Wen N, Peng D, Xiong X, et al. Cholangiocarcinoma combined with biliary obstruction: an exosomal circRNA signature for diagnosis and early recurrence monitoring. Signal transduction and targeted therapy 2024;9(1):107. doi: 10.1038/s41392-024-01814-3

15. Yang S, Fu J, Qin W, et al. Bile metabolic fingerprints distinguish biliary tract cancer from benign biliary diseases. Hepatology 2025;81(2):476–90. doi: 10.1097/hep.0000000000000957 [published Online First: 2024/06/11]

16. Arechederra M, Rullán M, Oyón D, et al. Bile as a liquid biopsy matrix: potential applications and limitations. Exploration of Digestive Diseases 2024;3(1):5–21.

17. Swanson K, Wu E, Zhang A, et al. From patterns to patients: Advances in clinical machine learning for cancer diagnosis, prognosis, and treatment. Cell 2023;186(8):1772–91. doi: 10.1016/j.cell.2023.01.035 [published Online First: 2023/03/12]

18. Haug CJ, Drazen JM. Artificial Intelligence and Machine Learning in Clinical Medicine, 2023. N Engl J Med 2023;388(13):1201–08. doi: 10.1056/NEJMra2302038 [published Online First: 2023/03/30]

19. From explanations to feature selection: assessing SHAP values as feature selection mechanism. 2020 33rd SIBGRAPI Conference on Graphics, Patterns and Images (SIBGRAPI); 2020 7-10 Nov. 2020.

20. Molnar C, Casalicchio G, Bischl B. iml: An R package for interpretable machine learning. Journal of Open Source Software 2018;3(26):786.

21. Abreu Lopez BA, Pinto-Colmenarez R, Caliwag FMC, et al. Colorectal Cancer Screening and Management in Low- and Middle-Income Countries and High-Income Countries: A Narrative Review. Cureus 2024;16(10):e70933. doi: 10.7759/cureus.70933 [published Online First: 2024/11/06]

22. Dangi RR, Sharma A, Vageriya V. Transforming Healthcare in Low-Resource Settings With Artificial Intelligence: Recent Developments and Outcomes. Public health nursing (Boston, Mass) 2025;42(2):1017–30. doi: 10.1111/phn.13500 [published Online First: 2024/12/04]

23. Björnsson E, Gustafsson J, Borkman J, et al. Fate of patients with obstructive jaundice. Journal of hospital medicine 2008;3(2):117–23. doi: 10.1002/jhm.272 [published Online First: 2008/04/29]

24. Siddique K, Ali Q, Mirza S, et al. Evaluation of the aetiological spectrum of obstructive jaundice. J Ayub Med Coll Abbottabad 2008;20(4):62–6. [published Online First: 2008/10/01]

25. Patel VB, Musa RK, Patel N, et al. Role of MRCP to determine the etiological spectrum, level and degree of biliary obstruction in obstructive jaundice. Journal of family medicine and primary care 2022;11(7):3436–41. doi: 10.4103/jfmpc.jfmpc_2362_21 [published Online First: 2022/11/18]

26. Garcea G, Ngu W, Neal CP, et al. Bilirubin levels predict malignancy in patients with obstructive jaundice. HPB (Oxford) 2011;13(6):426–30. doi: 10.1111/j.1477-2574.2011.00312.x [published Online First: 2011/05/26]

27. Chalya PL, Kanumba ES, McHembe M. Etiological spectrum and treatment outcome of Obstructive jaundice at a University teaching Hospital in northwestern Tanzania: A diagnostic and therapeutic challenges. BMC Res Notes 2011;4:147. doi: 10.1186/1756-0500-4-147 [published Online First: 2011/05/25]

28. Little JM, Cunningham P. Obstructive jaundice in a referral unit: surgical practice and risk factors. Aust N Z J Surg 1985;55(5):427–32. doi: 10.1111/j.1445-2197.1985.tb00917.x [published Online First: 1985/10/01]

29. Malchow-Møller A, Matzen P, Bjerregaard B, et al. Causes and characteristics of 500 consecutive cases of jaundice. Scand J Gastroenterol 1981;16(1):1–6. [published Online First: 1981/01/01]

30. Soto JA, Alvarez O, Lopera JE, et al. Biliary obstruction: findings at MR cholangiography and cross-sectional MR imaging. Radiographics 2000;20(2):353–66. doi: 10.1148/radiographics.20.2.g00mc06353 [published Online First: 2000/03/15]

31. Katabathina VS, Dasyam AK, Dasyam N, et al. Adult bile duct strictures: role of MR imaging and MR cholangiopancreatography in characterization. Radiographics 2014;34(3):565–86. doi: 10.1148/rg.343125211 [published Online First: 2014/05/14]

32. Sharma M, Pathak A, Sharma Y. Endoscopic ultrasound in CBD stone. Gastroenterology 2009;137(2):e7–8. doi: 10.1053/j.gastro.2009.01.059 [published Online First: 2009/07/02]

33. Lanzillotta M, Mancuso G, Della-Torre E. Advances in the diagnosis and management of IgG4 related disease. Bmj 2020;369:m1067. doi: 10.1136/bmj.m1067 [published Online First: 2020/06/18]

34. Okazaki K, Uchida K, Koyabu M, et al. IgG4 cholangiopathy: current concept, diagnosis, and pathogenesis. J Hepatol 2014;61(3):690–5. doi: 10.1016/j.jhep.2014.04.016 [published Online First: 2014/04/29]

35. Dyson JK, Beuers U, Jones DEJ, et al. Primary sclerosing cholangitis. Lancet (London, England) 2018;391(10139):2547–59. doi: 10.1016/s0140-6736(18)30300-3 [published Online First: 2018/02/18]

36. Lohse AW, Mieli-Vergani G. Autoimmune hepatitis. J Hepatol 2011;55(1):171–82. doi: 10.1016/j.jhep.2010.12.012 [published Online First: 2010/12/21]

37. Liu J, Qu J, Chen H, et al. The pathogenesis of renal injury in obstructive jaundice: A review of underlying mechanisms, inducible agents and therapeutic strategies. Pharmacological research 2021;163:105311. doi: 10.1016/j.phrs.2020.105311 [published Online First: 2020/11/28]

38. Martínez-Cecilia D, Reyes-Díaz M, Ruiz-Rabelo J, et al. Oxidative stress influence on renal dysfunction in patients with obstructive jaundice: A case and control prospective study. Redox biology 2016;8:160–4. doi: 10.1016/j.redox.2015.12.009 [published Online First: 2016/01/18]

39. Green J, Better OS. Systemic hypotension and renal failure in obstructive jaundice-mechanistic and therapeutic aspects. Journal of the American Society of Nephrology : JASN 1995;5(11):1853–71. doi: 10.1681/asn.v5111853 [published Online First: 1995/05/01]

40. Ilyas SI, Gores GJ. Pathogenesis, diagnosis, and management of cholangiocarcinoma. Gastroenterology 2013;145(6):1215–29. doi: 10.1053/j.gastro.2013.10.013 [published Online First: 2013/10/22]

41. van Oosten AF, Groot VP, Dorland G, et al. Dynamics of Serum CA19-9 in Patients Undergoing Pancreatic Cancer Resection. Ann Surg 2024;279(3):493–500. doi: 10.1097/sla.0000000000005977 [published Online First: 2023/06/30]

42. Liang B, Zhong L, He Q, et al. Diagnostic Accuracy of Serum CA19-9 in Patients with Cholangiocarcinoma: A Systematic Review and Meta-Analysis. Medical science monitor : international medical journal of experimental and clinical research 2015;21:3555–63. doi: 10.12659/msm.895040 [published Online First: 2015/11/19]

43. Loosen SH, Roderburg C, Kauertz KL, et al. CEA but not CA19-9 is an independent prognostic factor in patients undergoing resection of cholangiocarcinoma. Sci Rep 2017;7(1):16975. doi: 10.1038/s41598-017-17175-7 [published Online First: 2017/12/07]

44. Qin XL, Wang ZR, Shi JS, et al. Utility of serum CA19-9 in diagnosis of cholangiocarcinoma: in comparison with CEA. World J Gastroenterol 2004;10(3):427–32. doi: 10.3748/wjg.v10.i3.427 [published Online First: 2004/02/05]

45. Qurashi M, Vithayathil M, Khan SA. Epidemiology of cholangiocarcinoma. Eur J Surg Oncol 2025;51(2):107064. doi: 10.1016/j.ejso.2023.107064 [published Online First: 2023/09/15]

46. Fernandez YVM, Arvanitakis M. Early Diagnosis And Management Of Malignant Distal Biliary Obstruction: A Review On Current Recommendations And Guidelines. Clinical and experimental gastroenterology 2019;12:415–32. doi: 10.2147/ceg.s195714 [published Online First: 2019/12/07]

47. Podolsky DK, Camilleri M, Fitz JG, et al. Yamada’s textbook of gastroenterology: John Wiley & Sons 2015.

48. Yang K, Sun B, Zhang S, et al. RDW-SD is Superior to RDW-CV in Reflecting Liver Fibrosis Stage in Patients with Chronic Hepatitis B. Infection and drug resistance 2023;16:6881–91. doi: 10.2147/idr.s427047 [published Online First: 2023/11/03]

