## Supplementary material for "Integrating etiological insights with machine learning for precision diagnosis of obstructive jaundice: Findings from a high-volume center"

*Joint last author

^1^Division of Biliary Surgery, Department of General Surgery, West China Hospital, Sichuan University, Chengdu 610041, Sichuan, China

^2^Research Center for Biliary Diseases, West China Hospital, Sichuan University, Chengdu 610041, Sichuan, China

**Correspondence:**

Professor Nansheng Cheng

Postal address: Division of Biliary Surgery, Department of General Surgery, West China Hospital, Sichuan University, Chengdu 610041, Sichuan, China

or

Professor Jiong Lu

Postal address: Division of Biliary Surgery, Department of General Surgery, West China Hospital, Sichuan University, Chengdu 610041, Sichuan, China

or

Professor Bei Li

Postal address: Division of Biliary Surgery, Department of General Surgery, West China Hospital, Sichuan University, Chengdu 610041, Sichuan, China

or

Dr. Geng Liu

Postal address: Research Center for Biliary Diseases, West China Hospital, Sichuan University, Chengdu 610041, Sichuan, China

**Financial support statement:**

This work was supported by Sichuan Provincial Commission of Health Science Project (20PJ059); Sichuan Science and Technology Program (Grant No.2022YSF0060, Grant No.2022YSF0114, Grant No.2022NSFSC0680, Grant No. 2023YFS0094); 1·3·5 project for disciplines of excellence–Clinical Research Incubation Project, West China Hospital, Sichuan University (20HXFH021); 1·3·5 project for disciplines of excellence, West China Hospital, Sichuan University (ZYJC21049); The Key Research and Development Program sponsored by the Ministry of Science and Technology of Chengdu (Grant No. 2021-YF05- 00065-SN).

**Authors contributions:** Study concept and design: NW, YW, BL, JL, GL and NC; Data acquisition: NW, YW, YT, BL, JL and NC; Data analysis and interpretation: NW, YW, GL, JX and DZ; Implementation of machine learning: NW, YW, and GL; Drafting of the manuscript: NW, SW, GL, BL, JL and CN; Funding: XX, BL, JL and CN. All authors have read and critically revised the manuscript and agreed to the published version.

**Supplementary methods of data processing**

The following test results were included: α-fetoprotein (AFP, ng/mL), carcinoembryonic antigen (CEA, ng/mL), cancer antigen 125 (CA 125, U/mL), cancer antigen 19-9 (CA 19-9, U/mL), red blood cell count (RBC, ×10^12/L), hemoglobin (HGB, g/L), hematocrit (HCT, L/L), mean corpuscular volume (MCV, fL), mean corpuscular hemoglobin concentration (MCHC, g/L), mean corpuscular hemoglobin (MCH, pg), red cell distribution width-coefficient of variation (RDW-CV, %), red cell distribution width-standard deviation (RDW-SD, fL), platelet count (PLT, ×10^9/L), mean platelet volume (MPV, fL), platelet distribution width (PDW, %), large platelet ratio (P-LCR%, %), white blood cell count (WBC, ×10^9/L), neutrophil percentage (NEUT%, %), lymphocyte percentage (LYM%, %), monocyte percentage (MONO%, %), eosinophil percentage (EO%, %), basophil percentage (BASO%, %), neutrophil count (NEUT#, ×10^9/L), lymphocyte count (LYM#, ×10^9/L), monocyte count (MONO#, ×10^9/L), eosinophil count (EO#, ×10^9/L), basophil count (BASO#, ×10^9/L), total bilirubin (TBIL, μmol/L), direct bilirubin (DBIL, μmol/L), indirect bilirubin (IBIL, μmol/L), alanine aminotransferase (ALT, IU/L), aspartate aminotransferase (AST, IU/L), alkaline phosphatase (ALP, IU/L), γ-glutamyl transferase (γ-GT, IU/L), albumin (ALB, g/L), globulin (Glo, g/L), albumin/globulin ratio (A/G), glucose (GLU, mmol/L), urea (UREA, mmol/L), creatinine (CREA, μmol/L), cystatin C (CysC, mg/L), uric acid (UA, μmol/L), triglycerides (TG, mmol/L), cholesterol (CHOL, mmol/L), high-density lipoprotein cholesterol (HDL, mmol/L), low-density lipoprotein cholesterol (LDL, mmol/L), creatine kinase (CK, IU/L), lactate dehydrogenase (LDH, IU/L), hydroxybutyrate dehydrogenase (HBDH, IU/L), prothrombin time (PT, s), international normalized ratio (INR), activated partial thromboplastin time (APTT, s), fibrinogen (Fbg, g/L), thrombin time (TT, s), and C-reactive protein (CRP, mg/L).

Of note, one patient may undergo multiple examinations for the same item during the course of treatment. Only the laboratory results during the initial diagnosis of obstructive jaundice were utilized. If multiple test results still persist, median value was taken for further analysis.

**Supplementary methods of machine learning techniques**

The binary classification task focused on distinguishing between benign and malignant diseases. Various ML algorithms were utilized to achieve best predictive performance, including logistic regression (LR), decision tree models, K-nearest neighbors (KNN), random forest (RF), support vector machine (SVM), eXtreme Gradient Boosting (XGBoost), and lightGBM. The multi-class classification task intended to further categorize diseases into five detailed categories. These multi-class models were constructed using decision tree, XGBoost, RF, SVM, lightGBM, and KNN algorithms. All models were developed by R 4.1.3 (R Foundation for Statistical Computing, Vienna, Austria) using the mlr3 machine learning framework.

To assess the robustness and generalizability of the constructed models, internal validation and external validation were both conducted in binary classification models. Evaluation metrics included area under the receiver operating characteristic curve (AUROC) with 95% confidence intervals, accuracy (ACC), area under the precision-recall curve (AUPR), F1 score, sensitivity and specificity. Internal validation but not external validation was carried out for multi-class classification models, as the spectrum of diseases was limited in the BS cohort. Evaluation metrics of multi-class models included accuracy (ACC), area under the receiver operating characteristic curve weighted by prevalence (AUNU), Macro F1 score, precision score and recall score. Interpretability analysis was conducted on the optimized models with ‘iml’ and ‘mlr3verse’ in R. Feature importance score and SHapley Additive exPlanations (SHAP) values were calculated.

**Supplementary figures**

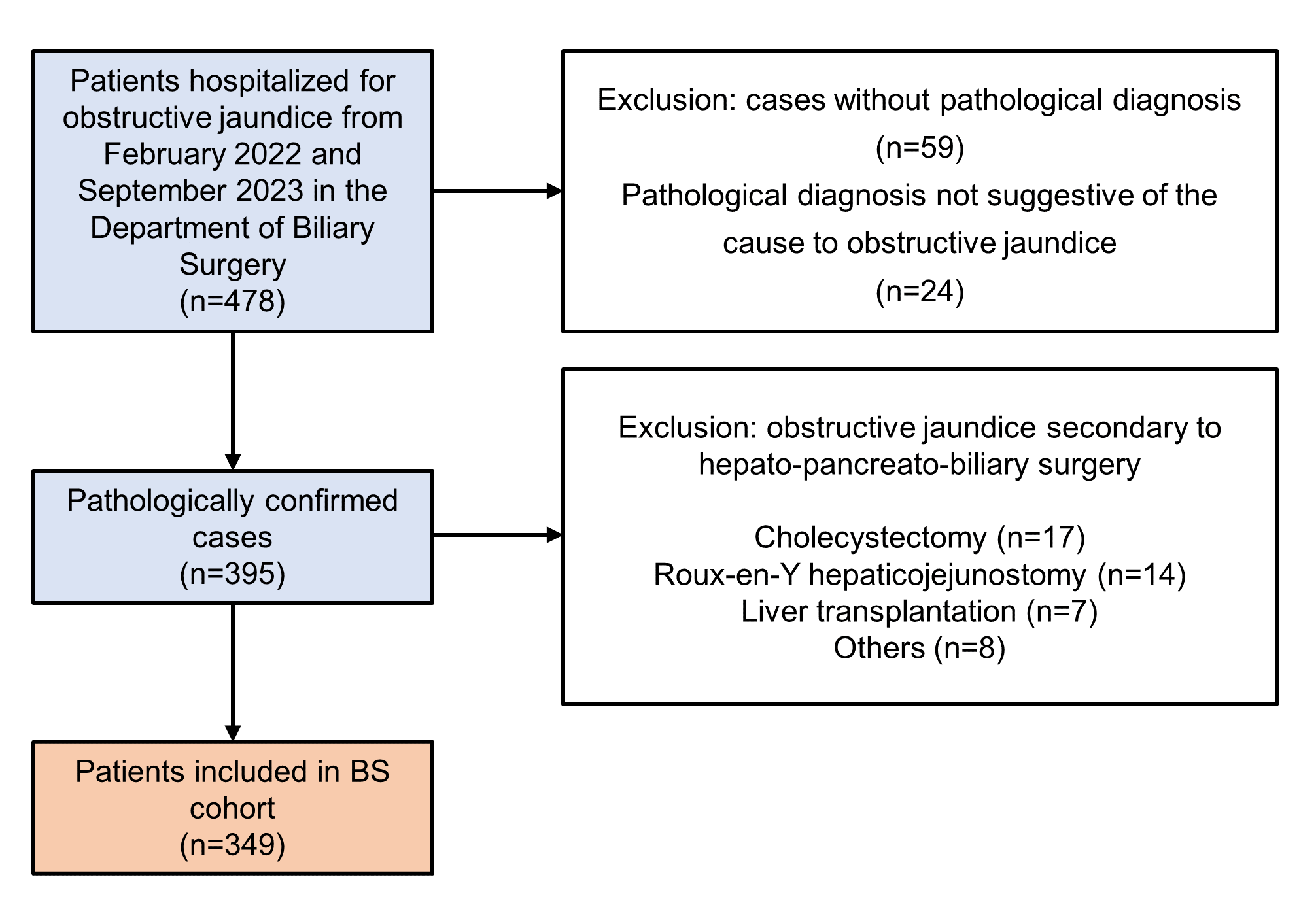

**Fig. S1 Flowchart illustrating the patient selection process to establish the BS cohort.**

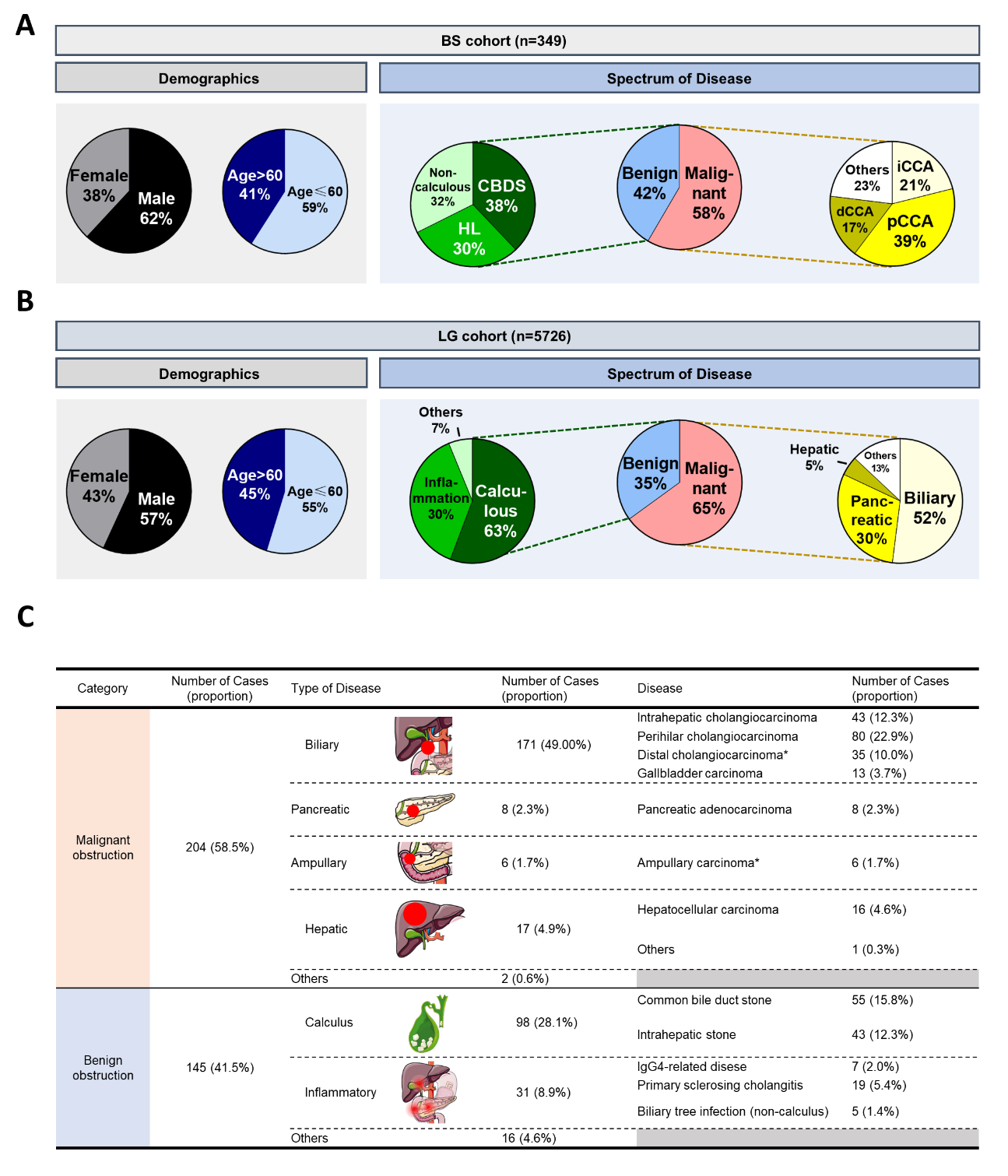

**Fig. S2 Summary of demographic characteristics and disease spectrum.** **(A)** Demographic characteristics and disease spectrum in the BS and LG cohorts were summarized in pie charts. **(B)** A more detailed investigation regarding disease spectrum in the BS cohort was also summarized.

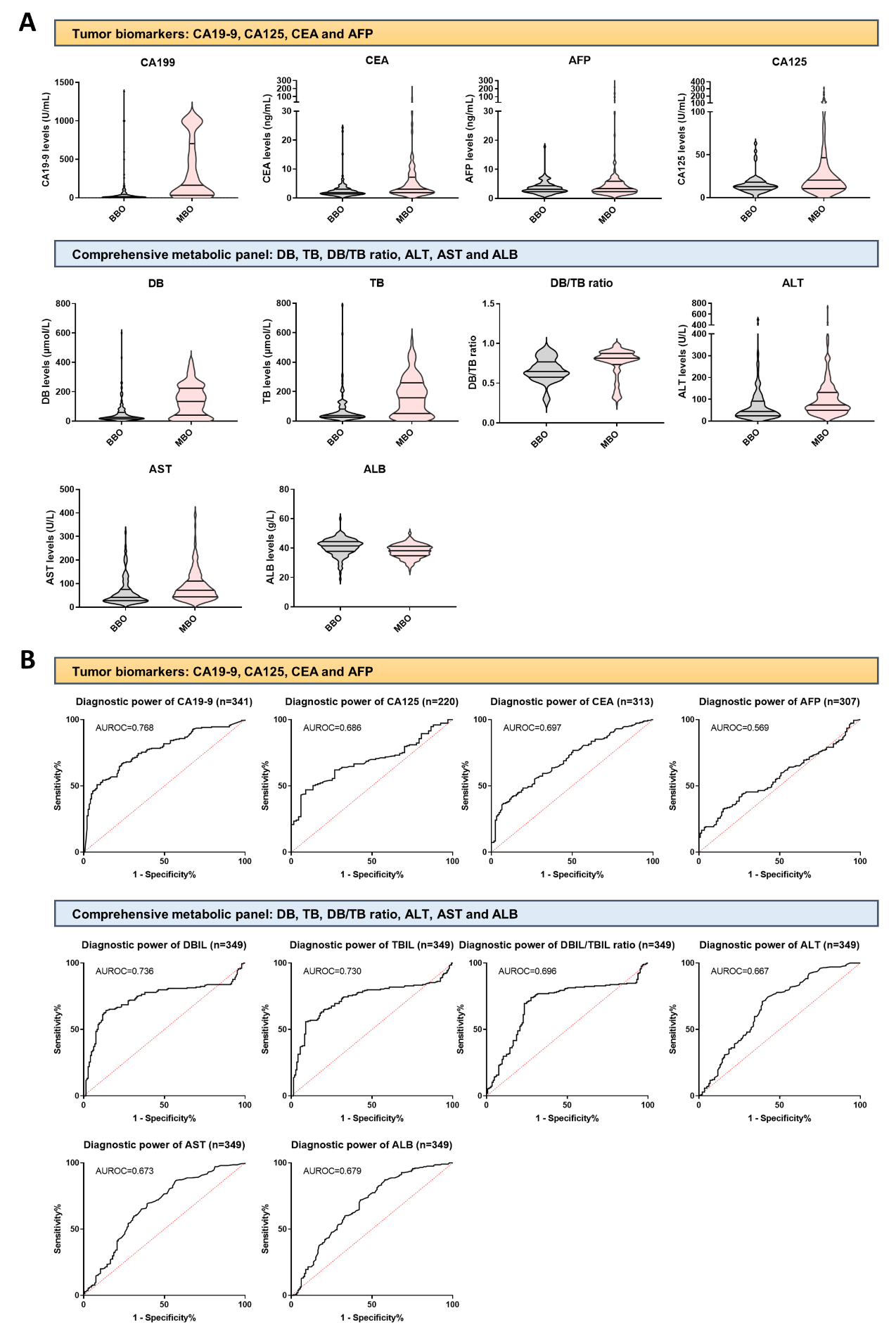

**Fig. S3 Potential and limitations of common clinical markers in the differential diagnosis of benign and malignant obstructions.** **(A)** Differences in clinical serum markers between benign and malignant groups in the BS cohort were visualized using violin plots. **(B)** Their efficacy as standalone diagnostic markers were also evaluated with ROC curves.

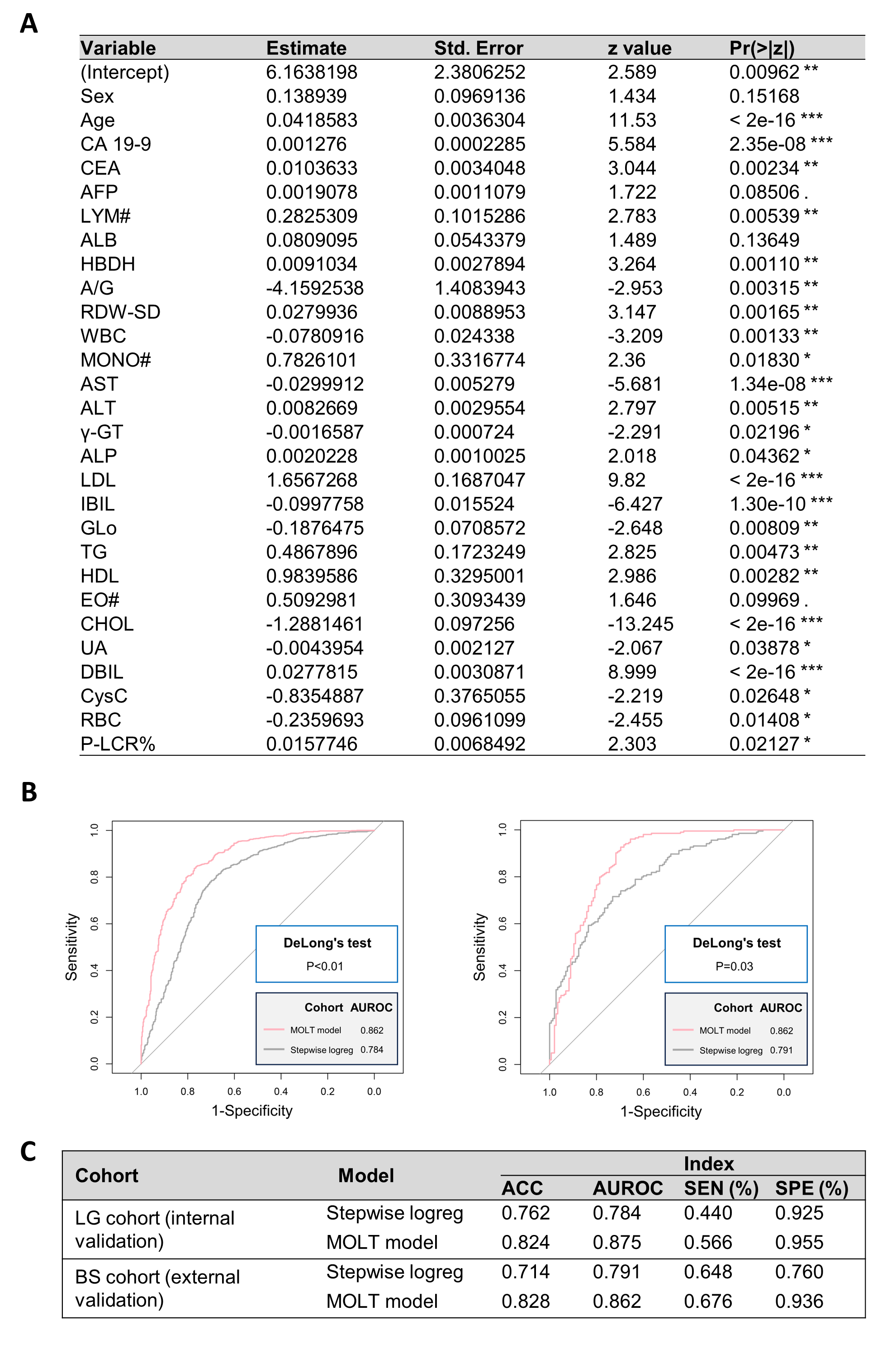

**Fig. S4 ML models outperformed stepwise logistic regression model. (A)** A stepwise logistic regression model differentiating benign from malignant obstructions was established via the training set in the LG cohort following inclusion and exclusion of variables from an initial pool of 57 indicators. **(B)** DeLong’s test revealed that this model had lower diagnostic power compared to the MOLT model we later established, both in the internal and external validation sets. **(C)** Other parameters for model evaluation also addressed this point.

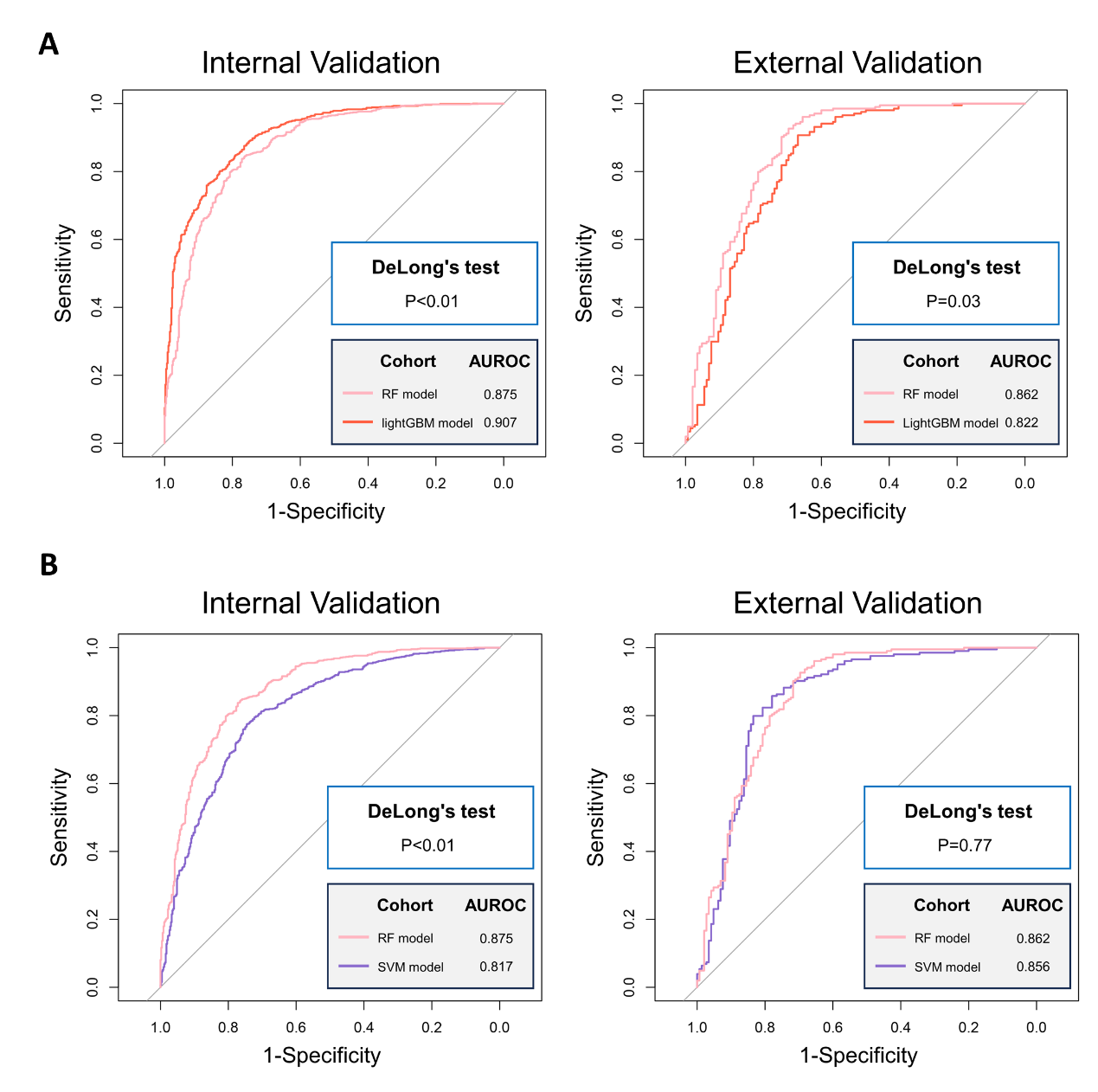

**Fig. S5 The RF model outperformed the others to be adopted as the MOLT model.** **(A)** Although the RF model had lower diagnostic power in the internal validation set compared with the lightGBM model, it stood out as the better one in the external validation set. **(B)** The SVM model, on the other hand, although exhibited comparable diagnostic power in the external validation set, was inferior to the RF model in the internal validation set.

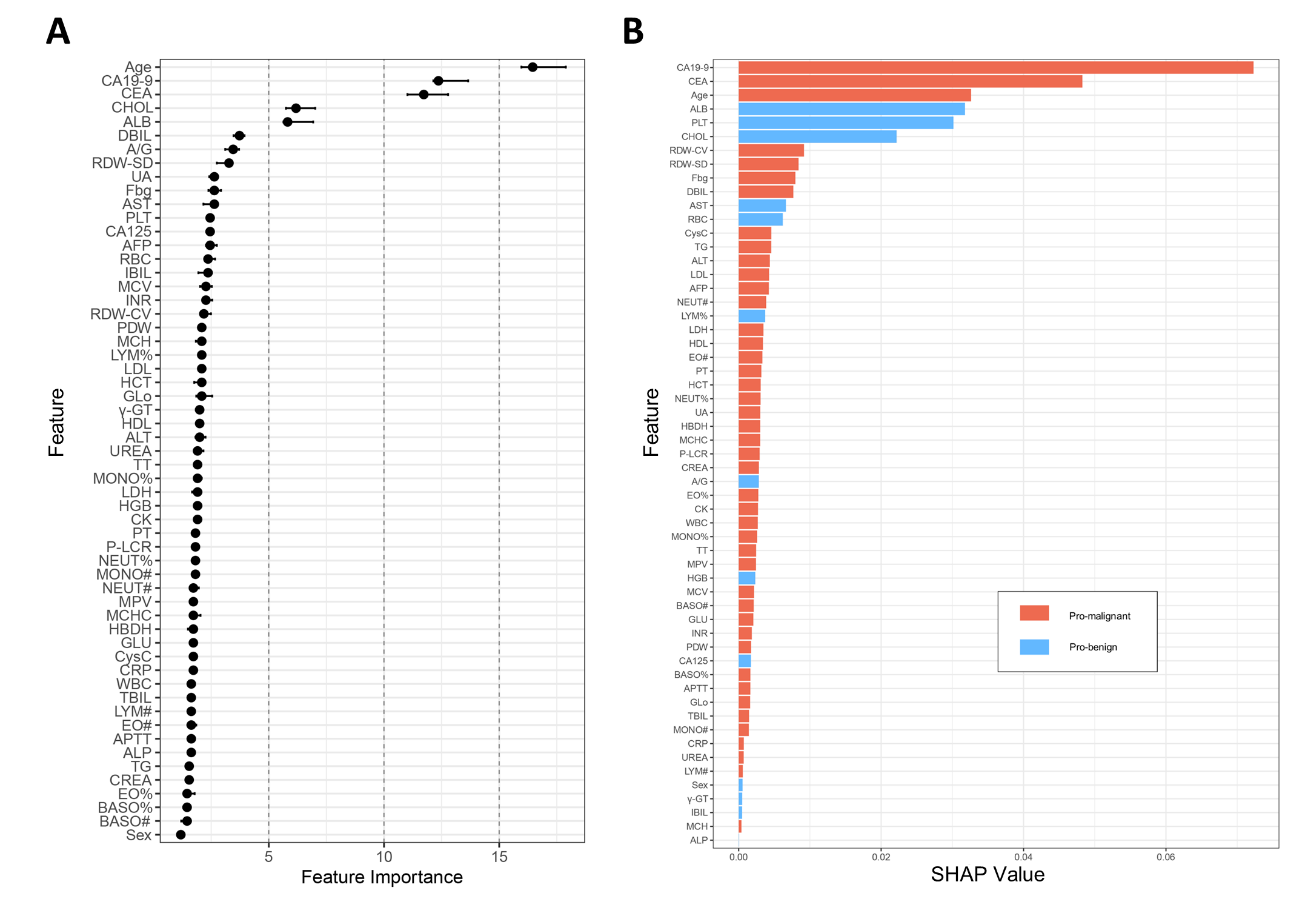

**Fig. S6 Interpretation of the binary MOLT model using feature importance score and SHAP value.** All 57 features were ranked by their **(A)** feature importance scores and **(B)** SHAP values to reveal their contribution to the binary MOLT model.

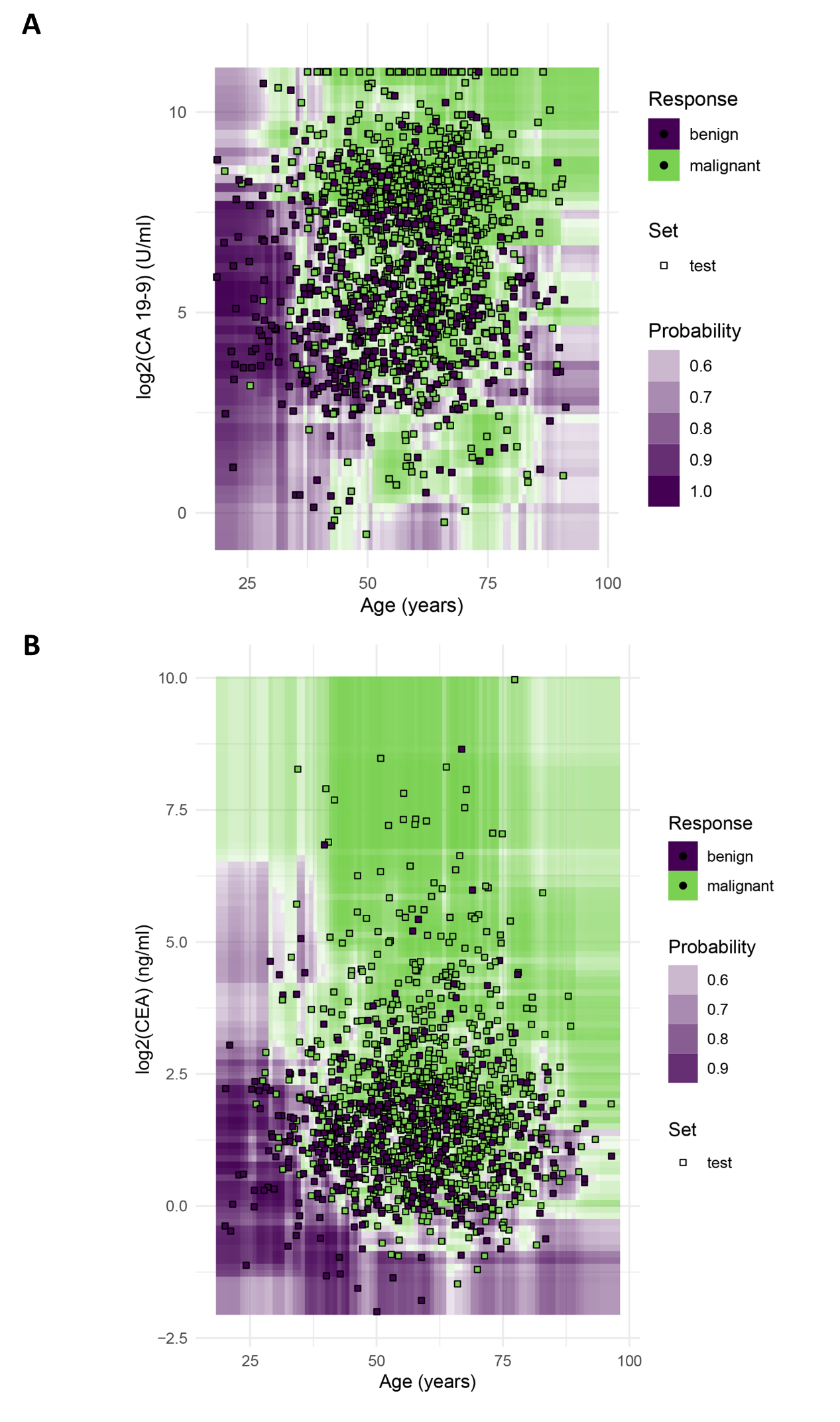

**Fig. S7 Visualization of decision boundaries pertaining to key features.** The decision boundaries of **(A)** CA 19-9 combining with age and **(B)** CEA combining with age were illustrated.

**Supplementary tables**

**Table. S1 Comparative analysis of baseline characteristics between the BS and LG cohorts.**

| Variables | BS cohort (n=349) | LG cohort (n=5726) | pVal |
| --- | --- | --- | --- |
| Category |  |  | 0.03 |
| Benign | 145 (41.5%) | 1994 (34.8%) |  |
| Borderline | 0 (0.0%) | 12 (0.2%) |  |
| Malignant | 204 (58.5%) | 3720 (65.0%) |  |
| Sex |  |  | 0.07 |
| Male | 216 (61.9%) | 3256 (56.9%) |  |
| Female | 133 (38.1%) | 2470 (43.1%) |  |
| Age (years) | 56.1 (11.8) | 57.8 (13.7) | 0.02 |
| CA 19-9 (U/ml) | 68.40 [14.30, 304.10] | 71.82 [23.23, 250.00] | 0.38 |
| CEA (ng/ml) | 2.44 [1.60, 4.55] | 2.90 [1.78, 5.16] | <0.01 |
| CA 125 (U/ml) | 15.27 [10.10, 27.00] | 22.60 [13.22, 48.00] | <0.01 |
| AFP (ng/ml) | 3.25 [2.33, 5.42] | 2.83 [2.05, 4.02] | <0.01 |
| TBIL (μmol/L) | 78.9 [62.6, 174.7] | 53.7 [43.6, 101.7] | <0.01 |
| DBIL (μmol/L) | 50.9 [41.2, 134.4] | 22.9 [15.6, 68.2] | <0.01 |
| IBIL (μmol/L) | 11.4 [8.3, 15.5] | 8.5 [6.1, 12.6] | <0.01 |
| ALT (IU/L) | 64 [35, 119] | 41 [26, 67] | <0.01 |
| AST (IU/L) | 58 [35, 102] | 37 [24, 61] | <0.01 |
| ALP (IU/L) | 144 [98, 262] | 134 [84, 227] | 0.01 |
| γ-GT (IU/L) | 203 [93, 440] | 93 [31, 234] | <0.01 |
| ALB (g/L) | 39.1 (5.3) | 37.9 (5.6) | <0.01 |
| Glo (g/L) | 27.8 (5.1) | 27.7 (5.6) | 0.78 |
| A/G | 1.34 [1.23, 1.45] | 1.40 [1.20, 1.58] | <0.01 |
| GLU (mmol/L) | 5.86 [5.52, 6.41] | 5.80 [5.20, 6.78] | 0.12 |
| UREA (mmol/L) | 4.8 (4.5, 5.2) | 4.7 (3.9, 5.7) | 0.08 |
| CREA (μmol/L) | 57.28 [54.72, 59.89] | 54.50 [49.00, 62.58] | <0.01 |
| CysC (mg/L) | 0.99 [0.91, 1.08] | 0.94 [0.84, 1.07] | <0.01 |
| UA (μmol/L) | 193 (48) | 214 (86) | <0.01 |
| TG (mmol/L) | 1.43 [1.25, 1.79] | 1.34 [1.02, 1.78] | <0.01 |
| CHOL (mmol/L) | 4.60 [4.20, 6.02] | 4.24 [3.42, 5.22] | <0.01 |
| HDL (mmol/L) | 0.76 [0.32, 0.99] | 0.92 [0.49, 1.31] | <0.01 |
| LDL (mmol/L) | 1.93 (0.81) | 2.10 (0.87) | <0.01 |
| CK (IU/L) | 71 [51, 86] | 68 [46, 102] | 0.99 |
| LDH (IU/L) | 187 [167, 198] | 178 [157, 201] | <0.01 |
| HBDH (IU/L) | 146 [129, 154] | 137 [123, 157] | <0.01 |
| RBC (×10^12/L) | 3.63 (0.64) | 3.67 (0.71) | 0.29 |
| HGB (g/L) | 136 (19) | 111 (20) | <0.01 |
| HCT (L/L) | 0.34 (0.06) | 0.34 (0.06) | 0.83 |
| MCV (fL) | 93.3 [90.3, 97.0] | 93.4 [89.9, 96.8] | 0.45 |
| MCHC (g/L) | 329 (11) | 327 (11) | <0.01 |
| MCH (pg) | 30.9 [29.7, 32.1] | 30.7 [29.4, 31.9] | 0.02 |
| RDW-CV (%) | 15.4 [14.4, 16.6] | 15.0 [13.9, 16.4] | <0.01 |
| RDW-SD (fL) | 51.8 [48.7, 56.1] | 50.7 [46.9, 55.2] | <0.01 |
| PLT (×10^9/L) | 160 [116, 205] | 192 [144, 244] | <0.01 |
| MPV (fL) | 12.1 (1.1) | 11.9 (1.2) | <0.01 |
| PDW (%) | 15.6 (3.0) | 15.2 (3.1) | <0.01 |
| P-LCR% (%) | 41.3 (9.0) | 39.6 (9.8) | <0.01 |
| WBC (×10^9/L) | 5.31 [4.31, 6.65] | 7.21 [5.68, 9.43] | <0.01 |
| NEUT% (%) | 72.5 [65.7, 79.3] | 73.1 [66.6, 79.1] | 0.34 |
| LYM% (%) | 16.4 [11.0, 22.4] | 15.8 [10.9, 22.0] | 0.25 |
| MONO% (%) | 7.0 [5.8, 8.3] | 7.1 [6.1, 8.4] | 0.09 |
| EO% (%) | 1.5 [0.9, 2.3] | 1.5 [0.9, 2.4] | 0.53 |
| BASO% (%) | 0.4 [0.2, 0.5] | 0.4 [0.3, 0.5] | 0.03 |
| NEUT# (×10^9/L) | 5.25 [3.64, 7.33] | 5.12 [3.76, 7.27] | 0.79 |
| LYM# (×10^9/L) | 1.15 [0.86, 1.50] | 1.11 [0.84, 1.45] | 0.14 |
| MONO# (×10^9/L) | 0.50 [0.40, 0.64] | 0.50 [0.40, 0.65] | 0.56 |
| EO# (×10^9/L) | 0.09 [0.06, 0.16] | 0.10 [0.06, 0.16] | 0.49 |
| BASO# (×10^9/L) | 0.03 [0.02, 0.04] | 0.03 [0.02, 0.04] | 0.07 |
| PT (s) | 11.6 [11.0, 12.3] | 11.7 [10.9, 12.7] | 0.05 |
| INR | 1.05 [0.98, 1.12] | 1.03 [0.97, 1.13] | 0.33 |
| APTT (s) | 28.4 [26.5, 31.0] | 28.1 [26.1, 30.8] | 0.37 |
| Fbg (g/L) | 3.53 (1.05) | 3.59 (1.14) | 0.32 |
| TT (s) | 19.0 [18.1, 20.0] | 18.2 [17.2, 19.4] | <0.01 |
| CRP (mg/L) | 51.2 [34.0, 67.5] | 17.1 [5.3, 59.7] | <0.01 |

Continuous variables with normal distribution are presented as mean value (SD) while others are presented as median [IQR]. Categorical variables are presented as frequency (percentage) unless otherwise stated.

**Table. S2 Comparative analysis of baseline characteristics between benign and malignant groups in the BS cohort.**

| Variables | BS cohort (n=349) | | |
| --- | --- | --- | --- |
|  | **Benign obstruction (n=145)** | **Malignant obstruction (n=204)** | **pVal** |
| Sex |  |  | <0.01 |
| Male | 77 (53.1) | 139 (68.1) |  |
| Female | 68 (46.9) | 65 (31.9) |  |
| Age (years) | 50.59 (12.91) | 59.95 (9.16) | <0.01 |
| CA 19-9 (U/ml) | 20.90 [7.26, 68.40] | 163.50 [34.80, 698.75] | <0.01 |
| CEA (ng/ml) | 1.84 [1.40, 3.14] | 3.00 [1.86, 6.95] | <0.01 |
| CA 125 (U/ml) | 12.88 [9.20, 18.10] | 19.45 [10.90, 46.60] | <0.01 |
| AFP (ng/ml) | 3.25 [2.31, 4.58] | 3.30 [2.34, 5.91] | 0.12 |
| TBIL (μmol/L) | 37.90 [25.60, 76.10] | 158.50 [52.40, 258.55] | <0.01 |
| DBIL (μmol/L) | 25.50 [16.10, 57.10] | 133.60 [41.45, 222.88] | <0.01 |
| IBIL (μmol/L) | 11.44 [9.59, 14.46] | 11.21 [7.69, 15.67] | 0.57 |
| ALT (IU/L) | 44.00 [24.00, 90.00] | 73.50 [49.00, 130.50] | <0.01 |
| AST (IU/L) | 42.00 [28.00, 75.00] | 72.00 [44.00, 111.00] | <0.01 |
| ALP (IU/L) | 149.74 [99.77, 264.42] | 142.02 [97.65, 259.94] | 0.82 |
| γ-GT (IU/L) | 202.51 [84.55, 481.25] | 204.22 [94.38, 382.91] | 0.84 |
| ALB (g/L) | 40.74 (5.78) | 37.85 (4.57) | <0.01 |
| Glo (g/L) | 28.60 (5.30) | 27.22 (4.93) | 0.01 |
| A/G | 1.34 [1.27, 1.43] | 1.35 [1.22, 1.47] | 0.72 |
| GLU (mmol/L) | 5.88 [5.66, 6.31] | 5.82 [5.43, 6.54] | 0.34 |
| UREA (mmol/L) | 4.87 [4.58, 5.16] | 4.82 [4.36, 5.30] | 0.40 |
| CREA (μmol/L) | 57.20 [55.00, 59.52] | 57.39 [54.44, 60.06] | 0.83 |
| CysC (mg/L) | 0.99 [0.93, 1.06] | 0.98 [0.90, 1.08] | 0.17 |
| UA (μmol/L) | 199.41 (34.53) | 188.93 (55.48) | 0.05 |
| TG (mmol/L) | 1.41 [1.30, 1.62] | 1.46 [1.21, 1.94] | 0.73 |
| CHOL (mmol/L) | 5.87 [4.45, 6.96] | 4.38 [3.62, 5.06] | <0.01 |
| HDL (mmol/L) | 0.83 [0.58, 0.97] | 0.66 [0.23, 1.02] | <0.01 |
| LDL (mmol/L) | 1.88 (0.72) | 1.97 (0.86) | 0.35 |
| CK (IU/L) | 72.53 [59.49, 81.35] | 70.54 [48.21, 90.34] | 0.68 |
| LDH (IU/L) | 188.06 [172.97, 196.43] | 185.26 [165.44, 200.66] | 0.44 |
| HBDH (IU/L) | 147.21 [130.20, 152.61] | 144.19 [127.55, 154.22] | 0.22 |
| RBC (×10^12/L) | 3.69 (0.67) | 3.59 (0.61) | 0.13 |
| HGB (g/L) | 134.52 (18.46) | 137.56 (19.25) | 0.14 |
| HCT (L/L) | 0.34 (0.06) | 0.34 (0.06) | 0.19 |
| MCV (fL) | 93.68 [90.08, 97.14] | 93.25 [90.54, 96.87] | 0.80 |
| MCHC (g/L) | 329.31 (11.56) | 329.04 (10.72) | 0.82 |
| MCH (pg) | 30.71 [29.65, 32.16] | 31.02 [29.80, 32.12] | 0.61 |
| RDW-CV (%) | 15.13 [13.98, 16.80] | 15.48 [14.52, 16.58] | 0.10 |
| RDW-SD (fL) | 51.21 [47.40, 55.70] | 52.27 [49.42, 56.32] | 0.09 |
| PLT (×10^9/L) | 164.83 [129.03, 207.47] | 156.26 [109.85, 202.85] | 0.03 |
| MPV (fL) | 11.96 (1.11) | 12.12 (1.15) | 0.22 |
| PDW (%) | 15.31 (2.90) | 15.84 (3.06) | 0.11 |
| P-LCR% (%) | 40.55 (8.80) | 41.79 (9.03) | 0.20 |
| WBC (×10^9/L) | 5.43 [4.40, 7.48] | 5.27 [4.27, 6.38] | 0.14 |
| NEUT% (%) | 70.21 [62.46, 77.44] | 74.31 [67.22, 80.04] | <0.01 |
| LYM% (%) | 18.14 [11.91, 24.87] | 14.91 [10.70, 20.90] | <0.01 |
| MONO% (%) | 7.17 [5.94, 8.50] | 6.90 [5.72, 8.22] | 0.11 |
| EO% (%) | 1.54 [0.91, 2.59] | 1.45 [0.83, 2.23] | 0.19 |
| BASO% (%) | 0.40 [0.20, 0.50] | 0.30 [0.20, 0.50] | 0.17 |
| NEUT# (×10^9/L) | 4.82 [3.73, 6.51] | 5.51 [3.64, 7.66] | 0.04 |
| LYM# (×10^9/L) | 1.18 [0.87, 1.62] | 1.12 [0.86, 1.42] | 0.16 |
| MONO# (×10^9/L) | 0.51 [0.40, 0.63] | 0.48 [0.40, 0.64] | 0.75 |
| EO# (×10^9/L) | 0.10 [0.06, 0.17] | 0.09 [0.06, 0.16] | 0.45 |
| BASO# (×10^9/L) | 0.03 [0.02, 0.04] | 0.03 [0.02, 0.04] | 0.35 |
| PT (s) | 11.40 [10.80, 12.20] | 11.70 [11.00, 12.30] | 0.02 |
| INR | 1.02 [0.97, 1.09] | 1.06 [1.00, 1.14] | <0.01 |
| APTT (s) | 28.20 [26.60, 30.90] | 28.50 [26.37, 31.02] | 0.49 |
| Fbg (g/L) | 3.45 (1.13) | 3.58 (0.99) | 0.26 |
| TT (s) | 18.82 [17.91, 19.98] | 19.08 [18.30, 20.02] | 0.12 |
| CRP (mg/L) | 46.10 [28.74, 62.19] | 53.95 [38.35, 72.58] | <0.01 |

Continuous variables with normal distribution are presented as mean value (SD) while others are presented as median (IQR). Categorical variables are presented as frequency (percentage) unless otherwise stated.
